# Systemic and mucosal IgA responses are variably induced in response to SARS-CoV-2 mRNA vaccination and are associated with protection against subsequent infection

**DOI:** 10.1101/2021.08.01.21261297

**Authors:** Salma Sheikh-Mohamed, Baweleta Isho, Gary Y.C. Chao, Michelle Zuo, Carmit Cohen, Yaniv Lustig, George R. Nahass, Rachel E. Salomon-Shulman, Grace Blacker, Mahya Fazel-Zarandi, Bhavisha Rathod, Karen Colwill, Alainna Jamal, Zhijie Li, Keelia Quin de Launay, Alyson Takaoka, Julia Garnham-Takaoka, Anjali Patel, Christine Fahim, Aimee Paterson, Angel Xinliu Li, Nazrana Haq, Shiva Barati, Lois Gilbert, Karen Green, Mohammad Mozafarihashjin, Philip Samaan, Patrick Budylowski, Walter L. Siqueira, Samira Mubareka, Mario Ostrowski, James M. Rini, Olga L. Rojas, Irving L. Weissman, Michal Caspi Tal, Allison McGeer, Gili Regev-Yochay, Sharon Straus, Anne-Claude Gingras, Jennifer L. Gommerman

## Abstract

Although SARS-CoV-2 infects the upper respiratory tract, we know little about the amount, type, and kinetics of antibodies (Ab) generated at this site in response to intramuscular COVID-19 vaccination, and whether these Ab protect against subsequent “breakthrough” infections. We collected longitudinal serum and saliva samples from participants receiving two doses of mRNA COVID-19 vaccines over a 6-month period and measured the relative level of anti-Spike and anti-Receptor Binding Domain (RBD) Ab. We detected anti-Spike/RBD IgG and IgA and associated secretory component in the saliva of most participants receiving 1 dose of mRNA vaccine. Administration of a second dose of mRNA boosted the IgG but not the IgA response, with only 30% of participants remaining positive for IgA at this timepoint. At 6 months post-dose 2, these participants exhibited greatly diminished anti-Spike/RBD IgG and IgA levels concomitant with a reduction in neutralizing activity in the saliva, although the level of secretory component associated anti-Spike was less susceptible to decay. Examining two prospective cohorts of subjects that were monitored for infections post-vaccination, we found that participants who were subsequently infected with SARS-CoV-2 had lower levels of vaccine-induced serum anti-Spike/RBD IgA at 2-4 weeks post-dose 2 compared to participants who did not experience an infection, whereas IgG levels were comparable between groups. These data emphasize the importance of developing COVID-19 vaccines that elicit a durable IgA response.

**One-Sentence Summary:** Our study delves into whether intra-muscular mRNA vaccination regimes confer a local IgA response in the oral cavity and whether the IgA response is associated with protection against breakthrough infection.

## Introduction

SARS-CoV-2 is a novel and highly contagious respiratory virus that has quickly spread across the globe. The virus uses a protein called Spike and its associated receptor binding domain (RBD) to interact with angiotensin converting enzyme 2 (ACE2) on the surface of epithelial cells in the upper respiratory tract^1^. SARS-CoV-2 is mainly transmitted via inhaled aerosols, making the immune response in the oral and nasal mucosa an important first line of defense^2^. Saliva is an important biofluid that can provide information about the mucosal antibody (Ab) response to SARS-CoV-2^3^. Indeed, the salivary glands themselves express ACE2 and harbor a significant population of IgA-producing plasma cells^3,4^. Secretory IgA (sIgA) in the saliva exists as IgA dimers that are associated with the secretory component, a proteolytic cleavage product which remains bound to IgA after it is transported across epithelial cells via the polymeric Ig receptor (pIgR)^5^. Secretory polymeric IgA has been shown to have potent neutralizing activity against SARS-CoV-2 *in vitro*^6^.

We and others have shown that IgM, IgG and IgA Ab against the SARS-CoV-2 Spike and RBD proteins are readily detected in the saliva of COVID-19 acute and convalescent patients^3,7^. Whether COVID-19 vaccines delivered through the parenteral intramuscular route (i.m.) generate a similar salivary antibody response is unclear, and the nature and kinetics of this response are ill-characterized. To address this gap, we measured SARS-CoV-2-specific Ab in saliva samples from participants who had received either BNT162b2 or mRNA-1273 vaccinations. We also determined whether levels of vaccine induced anti-Spike/RBD IgG or IgA differed in people who subsequently experienced a SARS-CoV-2 infection. Collectively, our data show that a durable IgA response is induced in approximately 30% of people vaccinated against COVID-19, and that IgA may play an important role in protection against infection.

## Materials and Methods

### Study Approvals

The Mount Sinai Hospital Research Ethics Board (REB) granted approval for recruiting staff in long-term care facilities located in the Greater Toronto Area for blood and saliva collection and for conducting serum ELISAs at the Lunenfeld-Tanenbaum Research Institute (study number: 20-0339-E). The University of Toronto REB granted approval for participant recruitment to collect blood and saliva samples and for conducting saliva ELISAs (study number: 23901). The University of Saskatchewan REB granted approval for saliva sample collection during the pre-COVID era (study number: BIO-USask-1579). The Sheba Medical Center REB granted approval for health care worker recruitment (study number: 8008-20-SMC).

### Recruitment and participants-LTCH cohort (Supplementary Table 1)

Staff working in Long-Term Care Homes (LTCHs) in Ontario were eligible to participate in the study if: (1) they were over the age of 18, (2) they were comfortable (a) reading and writing in English, (b) providing blood samples, and (c) sharing their COVID-19 diagnostic status with the study team. To invite LTCH staff to participate in the study, study staff approached the administrators and/or directors of LTCHs to assess if they were interested in being a participating site in the study. If they were interested, the administrators and/or directors shared information about the study with their staff and provided a deadline by which staff should opt-out if they did not want to be contacted about the study by a member of the study team. The LTCH administration then shared with the study team the contact information of all LTCH staff who did not opt-out of further communication about the study. The study team then contacted these staff by phone to invite them to participate in the study. This active recruitment strategy was paired with passive recruitment strategies, including having the participating LTCHs and/or relevant staff organizations share recruitment advertisements about the study, as well as having participants approach study staff when they were on-site for sample collection. A total of 12 LTCHs participated as a site in this study. Across these 12 sites, 647 individuals were invited to participate. Of these 647, a subset of individuals was not able to be reached by phone (n=242, 37%), were not eligible to participate (n=105, 16%), or refused to participate (n=131, 20%). Common reasons for participant ineligibility included being beyond 6 weeks after their 2^nd^ dose of their COVID-19 vaccine or not being interested in receiving a COVID-19 vaccine. The most common reason for refusal to participate was lack of interest in being part of research. Finally, a subset of individuals (n=13) withdrew prior to their first sample collection, resulting in a final sample size of 156 participants.

### Recruitment of negative control participants (Supplementary Table 2a)

Control saliva samples were collected from unexposed, asymptomatic individuals residing in an area of very low COVID-19 case numbers (Grey County, Ontario) and throughout the Greater Toronto Area (GTA) in April of 2020. Pre-COVID era samples were collected at the University of Saskatchewan. A pool of n=51 negative controls was used to generate a positive cutoff for all assays/

### Recruitment and participants-COVID-19 acute and convalescent participants (Supplementary Table 2b)

Acute and convalescent serum and saliva samples were obtained from patients identified by surveillance of COVID-19 (confirmed by PCR; in- and out-patients) by the Toronto Invasive Bacterial Diseases Network in metropolitan Toronto and the regional municipality of Peel in south-central Ontario, Canada (REB studies #20-044 Unity Health Network, #02-0118-U/05-0016-C, Mount Sinai Hospital). Consecutive consenting patients admitted to four TIBDN hospitals were enrolled: these patients had serum and saliva collected at hospital admission, and survivors were asked to submit repeat samples at 4-12 weeks PSO. Consecutive out-patients diagnosed at the same 4 hospitals prior to March 15^th^ and on a convenience sample of later days were approached for consent to collect serum and saliva at 4-12 weeks PSO. Patients were interviewed and patient charts reviewed to determine age, sex, symptom onset date, and disease severity (mild, moderate, and severe).

### Recruitment and participants – Medical Sciences Building (MSB). Supplementary Table 4

Pre-vaccination baseline, 2-3 weeks post-dose 1, 3 months post-dose 1- and 2-4-weeks post-dose 2 serum and saliva samples were obtained from an independently recruited cohort at the University of Toronto in Toronto, Ontario, Canada under REB protocol 23901. Of the n=31 recruited participants, 2 were excluded based on prior COVID-19 exposure and 2 left the study after the first sampling (moved out of country). For the 3 months post-dose 1, 11 out of the 27 eligible participants provided samples, whereas all other timepoints had n=27 participants. Upon arrival at the sampling site, participants gave informed consent, and serum and saliva samples were collected.

### Recruitment and participants –Long Term Care (LTCH) resident breakthrough cohort. Supplementary Table 5

Residents in this LTCH were offered mRNA-1273 on 4 January and 2 February 2021. The facility has resident rooms on the second through fifth floors and administrative/service areas in the basement and ground floors. By 28 March, 100 of 124 residents (81%) and 120 of 224 staff (54%) had received 2 vaccine doses, and 7 residents (6%) and 29 staff (13%) had received 1 dose. Ongoing pandemic measures included no visitors except essential caregivers, symptom surveillance twice daily for residents and staff/caregivers, staff/caregiver nasopharyngeal swab SARS-CoV-2 testing 2–3 times weekly by reverse-transcriptase polymerase chain reaction (rtPCR) or daily rapid antigen testing (Panbio), cohorting of staff by floor to limit any exposure to a single floor, and universal masking and eye protection for staff/caregivers. All positive rapid antigen tests were confirmed by SARS-CoV-2 PCR. All specimens PCR positive for SARS-CoV-2 were tested for N501Y and E484K mutations using a multiplex real-time PCR assay. Public Health Ontario Laboratory and/or Sunnybrook Health Sciences Centre performed whole-genome sequencing and assessed predicted Pango lineage, as described elsewhere^8^. On the 2 outbreak units, positive specimens with concentrations too low for mutation detection were assumed to be lineage P.1.

### Recruitment and participants - Sheba Health Care Workers breakthrough cohort. Supplementary Table 6

Beginning January 20, 2021, 11 days after the first staff members had received a second dose of the BNT162b2 vaccine, a study was initiated to identify every breakthrough infection, including asymptomatic infections. Serum analysed herein was collected at approximately 2-4 weeks post-dose 2. Data were collected until April 28, 2021. Efforts were extended to identify new cases with the use of daily health questionnaires, a telephone hotline, extensive epidemiologic investigations of exposure events, and contact tracing of infected patients and personnel. Testing for the presence of SARS-CoV-2 by RT-PCR was performed on fully vaccinated staff members who were symptomatic or had been exposed to an infected person, regardless of symptoms. Antigen-detecting rapid diagnostic testing (Ag-RDT) was available as an initial screening tool in the personnel clinic in combination with RT-PCR testing. A breakthrough infection was defined as the detection of SARS-CoV-2 on RT-PCR assay performed 11 or more days after receipt of a second dose of BNT162b2 if no explicit exposure or symptoms had been reported during the first 6 days. A matched case–control analysis was used to identify possible correlates of breakthrough infection. For the case–control analysis, control serum samples were selected from a prospective cohort study to analyze vaccine-induced immune responses and dynamics at the Sheba Medical Center. Each breakthrough case was matched with control samples that had been obtained from uninfected controls according to the following variables: sex, age, the interval between the second dose of BNT162b2 vaccine and serologic at the peri-infection timepoint, and immunosuppression status (see^9^. We compared antibody titers of cases versus controls obtained during the initial postvaccination period. Breakthrough cases for which serologic samples were not available were excluded from this analysis.

### Saliva collection

LTCH, MSB-1 and MSB-2 participants were told not to eat, drink or smoke at least 30 minutes prior to collection. Subsequently, saliva were collected using Salivette® tubes (Sarstedt, Numbrecht, Germany), a collection system which consists of a cotton ball which participants chew for exactly three minutes and place into a tube, which is then placed into a larger outer tube. The entire system is spun in a centrifuge at 1000 x g for five minutes at room temperature. The inner tube contains a hole at the bottom, which allows all the saliva absorbed by the cotton ball to filter into the larger outer tube. The total saliva volume from each participant was then separated into 300-500µl aliquots and stored at -80°C until the time of testing. Given that these samples were collected from vaccinated participants who reported no symptoms of COVID-19 infection, we did not conduct any measures for viral inactivation.

### Antigen production-Saliva assay

The expression, purification and biotinylation of the SARS-CoV-2 RBD and Spike ectodomain were performed as recently described^3^. The human codon optimized cDNA of the SARS-CoV-2 Spike protein was purchased from Genscript (MC_0101081). The soluble RBD (residues 328-528, RFPN…CGPK) was expressed as a fusion protein containing a C-terminal 6xHis tag followed by an AviTag. The soluble trimeric Spike protein ectodomain (residues 1-1211, MFVF…QYIK) was expressed with a C-terminal phage foldon trimerization motif followed by a 6xHis tag and an AviTag. To help stabilize the Spike trimer in its prefusion conformation, residues 682–685 (RRAR) were mutated to SSAS to remove the furin cleavage site and residues 986 and 987 (KV) were each mutated to a proline residue (***51***). Stably transfected FreeStyle 293-F cells secreting the RBD and soluble Spike trimer were generated using a previously reported piggyBac transposon-based mammalian cell expression system (***52***). Protein production was scaled up in 1L shake flasks containing 300 mL FreeStyle 293 medium. At a cell density of 10^6^ cells/mL, 1 μg/mL doxycycline and 1 μg/mL Aprotinin were added. Every other day 150 mL of medium was removed and replaced by fresh medium. The collected medium was centrifuged at 10000 × g to remove the cells and debris and the His-tagged proteins were purified by Ni-NTA chromatography. The eluted protein was stored in PBS containing 300 mM imidazole, 0.1% (v/v) protease inhibitor cocktail (Sigma, P-8849) and 40% glycerol at -12°C. Shortly before use, the RBD and Spike proteins were further purified by size-exclusion chromatography on a Superdex 200 Increase (GE healthcare) or Superose 6 Increase (GE healthcare) column, respectively. Purity was confirmed by SDS-PAGE. For the Spike protein, negative stain electron microscopy was used show evidence of high-quality trimers. The Avi-tagged proteins, at a concentration of 100 μM or less, were biotinylated in reaction mixtures containing 200 μM biotin, 500 μM ATP, 500 μM MgCl_2_, 30 μg/mL BirA, 0.1% (v/v) protease inhibitor cocktail. The mixture was incubated at 30°C for 2 hours followed by size-exclusion chromatography to remove unreacted biotin.

### Enzyme-linked immunosorbent assays for detecting anti-Spike and anti-RBD IgA, IgG and IgM in saliva

96-well plates pre-coated with streptavidin were used for all saliva assays. We have previously determined that coating plates with 50ul per well of 2ug/ml of biotinylated RBD or 20ug/ml of biotinylated Spike diluted in sterile phosphate-buffered saline (PBS) was the ideal coating solution. Control wells were coated with 50ul per well of sterile PBS. After coating with the antigen of interest and incubating overnight at 4°C, the coating solution was discarded and plates were blocked with 5% BLOTTO solution (5% w/vol skim milk powder (BioShop, CAT# SKI400.500)). Plates were incubated with the blocking solution at 37°C for 2 hours, and the solution was discarded immediately prior to adding samples to each well. During the blocking incubation, frozen saliva samples were removed from -80°C storage, thawed and diluted using 2.5% BLOTTO at a range of 1:5-1:20. Sample dilutions were pre-incubated in a separate streptavidin-coated plate with no antigen to reduce anti-streptavidin activity in the saliva. Samples were incubated in the pre-adsorption plate for 30 minutes at 37°C, after which 50ul of each sample from the pre-adsorption plate was transferred to the corresponding wells of the antigen-coated plates and incubated for 2 hours at 37°C. Next, samples were discarded from the antigen-coated plates, and the plates were washed 3x with PBS+0.05% Tween 20 (PBS-T (BioShop, CAT# TWN510)). 50ul of Horse radish peroxidase (HRP)-conjugated goat anti human-IgG, IgA and IgM secondary antibodies (Southern Biotech, IgG: 2044-05, IgA: 2053-05, IgM: 2023-05) were added to the appropriate wells at dilutions of 1:1000, 1:2000 and 1:1000 in 2.5% BLOTTO, respectively, and incubated for 1 hour at 37°C. Plate development was done by adding 50uL of 3,3’,5,5’tetramethylbenzidine (TMB) Substrate Solution (ThermoFisher, 00-4021-56) to each well. The reaction was then stopped by adding 50µl/well of 1N H_2_SO_4_, and optical density (OD) was read at a wavelength of 450nm (OD_450_) on a spectrophotometer (Thermo Multiskan FC).

### ELISA for detection of secretory component associated anti-Spike/RBD antibodies

Secretory chain associated antibodies were detected by modifying our saliva Spike/RBD ELISA by using an HRP-conjugated Goat anti-human secretory component detection reagent at a dilution of 1/750 from Nordic MUBio (Cat# GAHu/SC/PO).

### Enzyme-linked immunosorbent assays for detecting anti-Spike and anti-RBD IgA, IgG and IgM in serum

Briefly, an automated chemiluminescent ELISA assay was used to analyze the levels of IgG, IgA, and IgM antibodies to the Spike trimer, and its RBD with the following modifications: RBD (PRO1151, 20 ng/well) antigens were produced in CHO (Chinese Hamster Ovary) cells, and were a kind gift from Dr. Yves Durocher, National Research Council of Canada (NRC). The secondary antibody for IgG was an IgG-HRP fusion (PRO1146, 1:6700 or 0.9 ng/well), donated by the NRC. A standard curve of the VHH72 monoclonal antibody^10^ fused to a human IgG1 Fc domain (PRO23, also from the NRC) was generated for calibrating the anti-Spike and anti-RBD IgG response. All other antigens, detection reagents and calibration reagents were as previously described^3^. The data analysis also proceeded as in^3^, with the following exceptions: Blanks were not subtracted from the chemiluminescence raw values of the samples, and the raw values were normalized to a blank-subtracted point in the linear range of the calibration standard curve (for Spike and RBD, the reference point was 0.0156 µg/ml and for N either 0.0625 µg/ml or a combination of 0.0156 and 0.0312 ug/ml was used to maintain consistent values across tests that used different anti-N antibody batches). The results are represented as a “relative ratio” to this reference point. To define the cutoff for positive antibody calls for each antigen for IgG when 0.0625 µl/well of sample was added, 3 standard deviations from the mean of the log negative control distribution from >20 different runs collected over 4 months was used. For IgA, negatives from 2 different runs over one month and for IgM negatives from 3 runs over 2 months were used. In all cases, the selected cut offs correspond to <2% False Positive Rate (FPR) assessment, based on Receiver Operating Characteristic Curves. For each sample, the raw OD_450_ for the PBS control well was subtracted from the raw antigen-specific OD_450_ value for each sample, at each sample dilution (1:5, 1:10, 1:20). The adjusted OD_450_ value for each sample dilution (1:5, 1:10, 1:20) was used to calculate the area under the curve (AUC) for each individual sample. The sample AUC was then normalized to the AUC of the positive control, which consisted of saliva collected from COVID-19 acute and convalescent participants. The normalized AUC was multiplied by 100 to give a final percentage, which we deemed the “% of positive control”. Each plate also included 1-3 negative controls (with a total of n=51 negative controls used), which consisted of pre-COVID era saliva incubated in antigen-coated wells. Integrated scores were calculated for all negative control samples, using the same calculation method used for cohort 1 and 2 samples. “Positive” cut off values for each antigen-specific isotype were calculated using the following formula: average integrated scores of negative samples + 2(standard deviation of negative control integrated scores). Samples whose score was above the resulting cut off for each antigen-specific isotype was used to determine which samples from cohorts 1 and 2 had detectable antibody levels in their saliva.

### Flow Cytometry method for detection of neutralizing activity

Neutralizing activity was measured at two step saliva dilutions (1:12-1:384) following incubation with recombinant Vesicular Stomatitis Virus (rVSV)-eGFP-SARS-CoV-2-Spike in which the VSV-G protein was replaced with SARS-CoV-2-Spike protein was propagated on MA104 cells^11^. MA104 cells were maintained in Medium 199 (Gibco, Cat. No. 11150067) supplemented with 10% FBS and 1% Penicillin/Streptomycin (Fisher Scientific, Cat. No. 15-140-163). After visible cytopathic affect, supernatant was filtered, aliquoted and stored at –80 °C. Supernatant was added to Human Embryonic Kidney (HEK) 293 cells were engineered to encode human angiotensin converting enzyme 2 (hACE2 in the pDEST-mCherry vector) as previously described^12^. HEK293-hACE2-mCherry cells were cultured in Gibco Dulbecco’s Modified Eagle Medium (DMEM) formulation containing glucose, L-glutamine and sodium pyruvate (Gibco, Cat. No. 11995065) with 1x Penicillin/Streptomycin. Geneticin Selective Antibiotic (G418) (Gibco, Cat. No. 10131035) was added at a concentration of 500 u μ/mL to maintain hACE2-mCherry expression. Cells were grown in 5% carbon dioxide (CO2) at 37 °C and passaged every 3 days using Versene solution (Gibco, Cat. No. 15040066). HEK293-hACE2-mCherry cells were seeded at a density of 25,000 cells per well in a 96-well, flat-bottom tissue culture coated plate. Outer rows were avoided to reduce assay variations resulting from edge effect in the IncuCyte. In a separate 96 well plate, samples were serially diluted and incubated with 50 μl of rVSV-eGFP-SARS-CoV-2-S for 2 hours at 37 °C in 5% CO2. Each sample plate included a dilution of anti-RBD antibody (Invitrogen, Cat. No. 703958) of 10 μg/mL, 5 μg/mL, 1 μg/mL, 0.5 μg/mL, 0.1 μg/mL, and 0.05 μg/mL as a positive control. After incubation, the mixture of sample and rVSV-eGFP-SARS-CoV-2-S was transferred to the plated HEK293-hACE2-mCherry cells at a 1:1 ratio of culture media to virus/sample suspension. Plates loaded in the IncuCyte were imaged every 1-4 hours for a total of 72 hours with 4 scans per sample well to visualize neutralization. The total integrated intensity of the fluorescent value of the lowest anti-RBD condition (0.05 μg/mL) with the rVSV-eGFP-SARS-CoV-2-S controls at 12-hour intervals was used to normalize separate experiment runs. Each plate was normalized either to the mean of the rVSV-eGFP-SARS-CoV-2-S supernatant controls or to the 0.05 μg/mL anti-RBD antibody. Normalization to 0.05 μg/mL anti-RBD was performed only if division by the triplicate rVSV-eGFP-SARS-CoV-2 control conditions resulted in loss of a sigmoidal shape of the anti-RBD curve. To quantitatively determine assay sensitivity, fluorescent reduction of neutralization titers (FRNT) 50 and 70 were calculated to determine the amount of monoclonal anti-RBD needed to prevent 50% and 30%, respectively, of the maximum infection (Sup. Fig. 10B and C) and plotted in log10 scale for plasma and log2 scale for saliva due to variations in antibody titers in different tissue types. This assay is sufficiently sensitive down to 5 μg/mL of neutralizing antibodies.

### Statistics

Antibody levels from the cross-sectional LTCH cohort were compared between groups using Mann Whitney U tests. For correlation analyses, a nonparametric Spearman correlation test was applied. A Friedman pair-wise multiple comparison test was used to compare participants who were assessed at multiple timepoints (Fig. 2), whereas the Wilcoxon pair-wise signed-rank test was used when examining 2 different timepoints (Fig. 4, Supplemental Fig. 5). Kruskal-Wallis with Dunn’s multiple comparison test was conducted to examine the differences in antibody levels to RBD and Spike between infected vs non-infected participants in the LTCH and Sheba cohorts. Adjusted P value based on 2 comparisons per family (RBD case vs. control and Spike case vs. control) were used to determine statistical significance. Analyses were performed in Prism (GraphPad) Version 9.2.0.

### Multivariable analysis (Supplementary Table 3a-c)

The relationships between clinical predictors (age, sex, SARS-CoV-2 infection prior to vaccination, and time from vaccination to sample collection) and antibody levels were examined in bivariate and *a priori* multivariable linear regression models. For saliva, four multivariable linear regression models were constructed to examine potential independent associations between the four clinical predictors and anti-RBD IgA/IgG and anti-Spike IgA/IgG. Kaplan-Meier survival curve were plotted by dividing each group based on serum titre at above or below median. Log-rank (Mantel-Cox) test was done to compare survival curve of each isotype (IgG and IgA anti-Spike) above and below median cohort values.

## Results

### Detection of anti-Spike and anti-RBD antibodies in saliva from participants receiving COVID-19 mRNA vaccines

We first compared saliva from long-term care home (LTCH) workers that received either BNT162b2 or mRNA-1273 (Supplementary Fig. 1 and Supplementary Table 1) with pooled negative control saliva used to establish a cut-off (Supplementary Tables 2a-b), and saliva from COVID-19 acute and convalescent patients as positive controls (Supplementary Table 2c). We expressed the data as a percentage relative to a pooled sample of saliva from COVID-19 acute and convalescent patients – the same pooled sample was present in each plate. We previously found that this method provided excellent plate-to-plate consistency and produced similar results as what we found when we normalized to total IgM/IgG/IgA^3^. After two doses of mRNA vaccine, 11%, 94% and 41% of participants were positive for anti-Spike IgM, IgG and IgA and 22%, 93% and 20% of participants were positive for anti-RBD IgM, IgG and IgA Ab (Fig. 1). Furthermore, as we observed before in COVID-19 recovered patients^3^, levels of salivary anti-Spike/RBD Ab correlated well with anti-Spike/RBD Ab in the serum (Supplementary Fig. 2). In multivariable analysis, age and prior SARS-CoV-2 infection were independently associated with the salivary anti-Spike IgA response (Supplementary Table 3a). In contrast, male sex had a negative independent association with the salivary anti-Spike IgG response (Supplementary Table 3b) as has been observed before for COVID-19 and other vaccines^13,14^. Lastly, prior SARS-CoV-2 infection and time since vaccination were independently associated with higher and lower serum anti-RBD IgA levels, respectively (Supplementary Table 3c).

**Fig 1.**
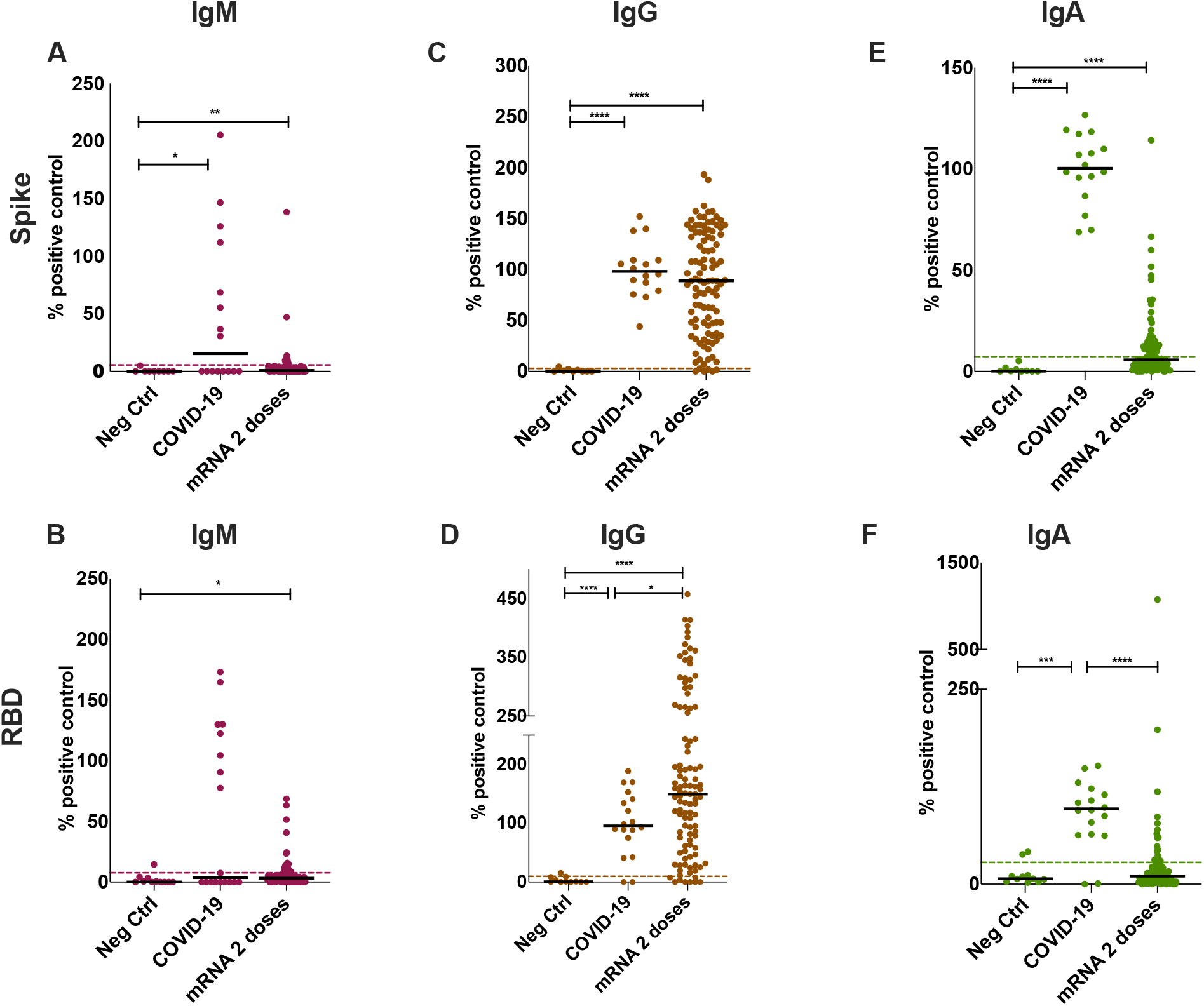
Analysis of anti-Spike and anti-RBD antibodies in saliva from LTCH workers receiving mRNA COVID-19 vaccines. Anti-Spike (**A,C,E)** and anti-RBD (**B,D,F**) antibodies were detected using an ELISA-based assay in the saliva of vaccinated LTCH workers after two-doses of either BNT162b2 or mRNA-1273. COVID-19 controls consisted of saliva collected from acute and convalescent patients (n = 18). These were compared to n=9 individually run negative controls. The positive cutoff (dotted line) was calculated as 2 standard deviations above the mean of a pool of negative control samples (n=51) for each individual assay. All data is expressed as a percentage of the pooled positive plate control, calculated using the AUC for each sample normalized to the AUC of the positive control (see Methods). Solid black bars denote the median for each cohort. Mann-Whitney U test was used to calculate significance, where *=p<0.05, **=p<0.01, ***=p<0.001 and ****=p<0.0001.

**Fig 2.**
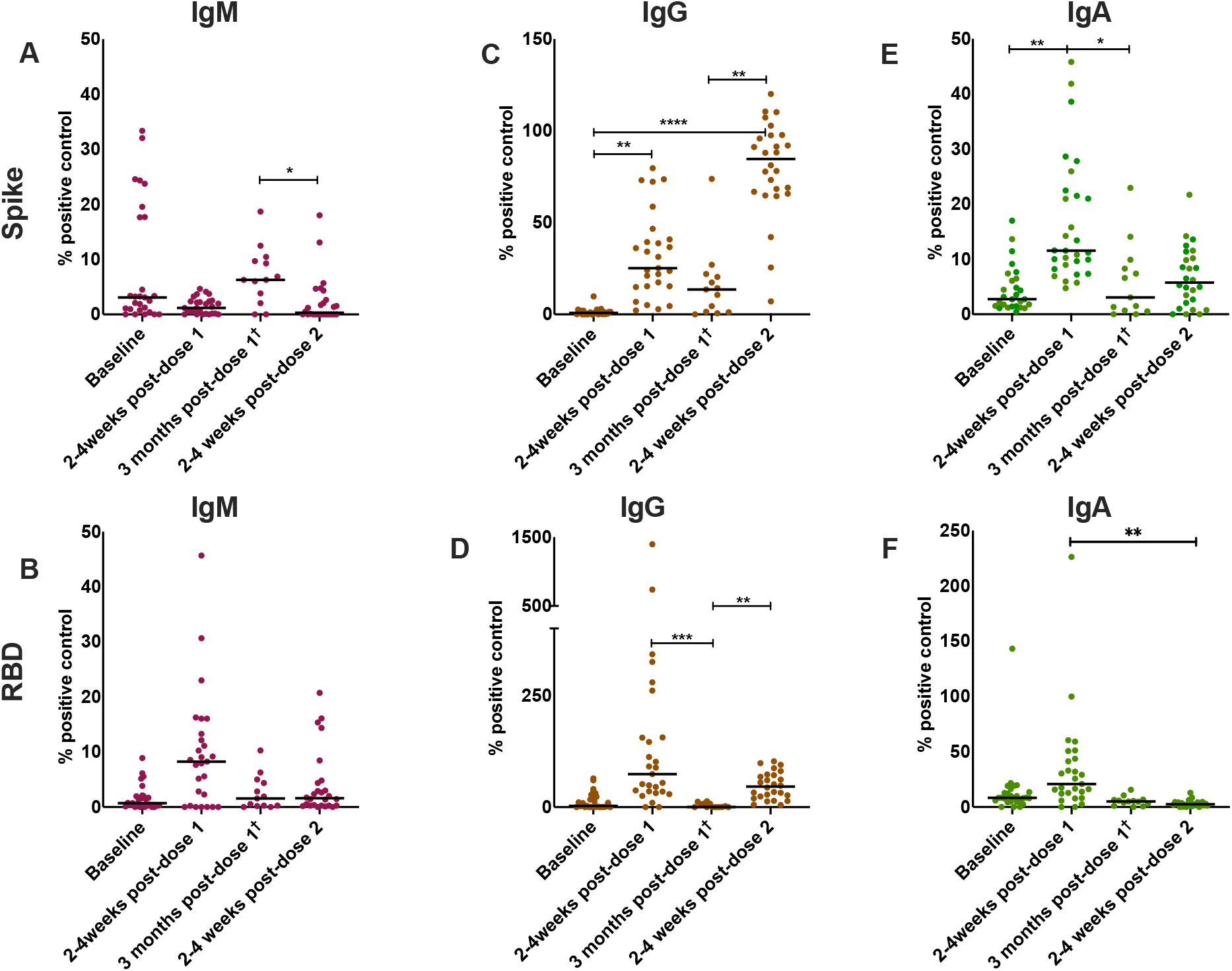
Analysis of anti-Spike and anti-RBD antibodies in saliva from sequentially sampled participants receiving COVID-19 mRNA vaccines. Anti-Spike (**A,C,E**) and anti-RBD (**B,D,F**) antibodies were analyzed in the saliva of vaccinated participants that were followed at sequential timepoints before and after dose 1 and dose 2 of BNT162b2 (MSB-1, n=27 with † = n=11 participants at this timepoint). All data is expressed as a percentage of the pooled positive plate control, calculated using the AUC for each sample normalized to the AUC of the positive control (see Methods). Solid black bars denote the median for each cohort. A Friedman pair-wise multiple comparison test was used for pair-wise comparisons for multiple timepoints, considering only data points from participants who had completed all timepoints (n=11). *=p<0.05, **=p<0.01, ***=p<0.001 and ****=p<0.0001.

### Longitudinal assessment of anti-Spike and anti-RBD antibodies in saliva from participants receiving 2 doses of COVID-19 mRNA vaccines at a 3 month dose interval

As of June 2021, although most LTCH workers had been fully vaccinated, significant sectors of the Canadian population had only been administered a single dose of a COVID-19 vaccine because the interval between dose 1 vs dose 2 was extended as a dose sparing measure. Thus, we wished to ascertain if salivary Ab could be detected after a single dose of a COVID-19 mRNA vaccine, and how long these Ab would persist. We therefore collected samples from a second cohort of healthy adults that were followed longitudinally (Medical Sciences Building cohort – MSB; Fig. 1 and Supplementary Table 4). These participants received 1 dose of BNT1162b2 and a second dose of BNT1162b2 3 months later, with samples taken at baseline, 2 weeks post-dose 1, 3 months post-dose 1, and 2-weeks post-dose 2. For the entire sampling period, of those participants that did not show evidence of prior SARS-CoV-2 infection IgM levels were higher than baseline only for anti-Spike post-dose 2, and anti-RBD at 2 weeks post-dose 1. Focusing therefore on IgG and IgA, we observed that 97% and 93% of participants were positive for anti-Spike IgG and IgA, and 52% and 41% were positive for anti-RBD IgG and IgA Ab in their saliva 2 weeks post-dose 1 (Fig. 2). Of note, three months after dose 1, the median level of salivary anti-Spike/RBD Ab had diminished nearly to baseline. Following administration of a second dose of mRNA, while antigen-specific IgG levels recovered upon administration of dose 2 as expected, a second dose did not further augment antigen-specific salivary IgA levels in most subjects, and only approximately 30% of participants remained positive for IgA after dose 2 (Fig. 2C-D vs 2E-F). Therefore, the IgG and IgA response to COVID-19 mRNA vaccination differs upon administration of dose 2.

### Detection of secretory component associated with anti-Spike and anti-RBD antibodies in saliva from participants receiving COVID-19 mRNA vaccines

The i.m. route induces an immune response in the axillary draining lymph node that is biased towards class switch to IgG rather than IgA. Thus, we were curious as to why we were able to detect an IgA anti-Spike/RBD response in the saliva, particularly after one dose of mRNA vaccine. We hypothesized that some IgA may be produced locally. To test this, we designed an ELISA to detect secretory chain associated with anti-Spike/RBD Ab. We determined that the secretory chain signal associated with anti-SARS-CoV-2 salivary Ab could be out-competed with recombinant secretory chain, and no anti-Spike/RBD secretory chain signal was detected in pre-pandemic colostrum, demonstrating assay specificity (Supplementary Fig. 3). We then measured secretory component associated anti-SARS-CoV-2 Ab in the saliva of vaccinated LTCH participants who had received two doses of either BNT162b2 or mRNA-1273. We found that secretory component associated anti-Spike and anti-RBD Ab could be detected in 30% and 58% of participants, respectively, although the levels were significantly lower than what was observed in COVID-19 patients (Fig. 3A-B). The anti-SARS-CoV2 associated secretory chain signal was independent of prior SARS-CoV-2 exposure as we observed no significant difference in this signal comparing participants who were positive vs negative for serum anti-nucleocapsid protein Ab (Fig. 3C-D). Of note, if we divided the LTCH cohort into those participants who were positive versus negative for anti-Spike/RBD IgA, we observed that the secretory component signal was only detected in the IgA^+^ participants (Fig. 3E-F). Moreover, anti-Spike/RBD IgA and the level of secretory component positively correlated with each other at (Supplementary Fig. 4). Combined with the finding that most participants did not produce IgM Ab to anti-Spike/RBD, we conclude that secretory component is associating with anti-Spike/RBD IgA (sIgA). Therefore, a local sIgA response to Spike/RBD is produced in response to mRNA vaccination in some participants.

**Fig 3.**
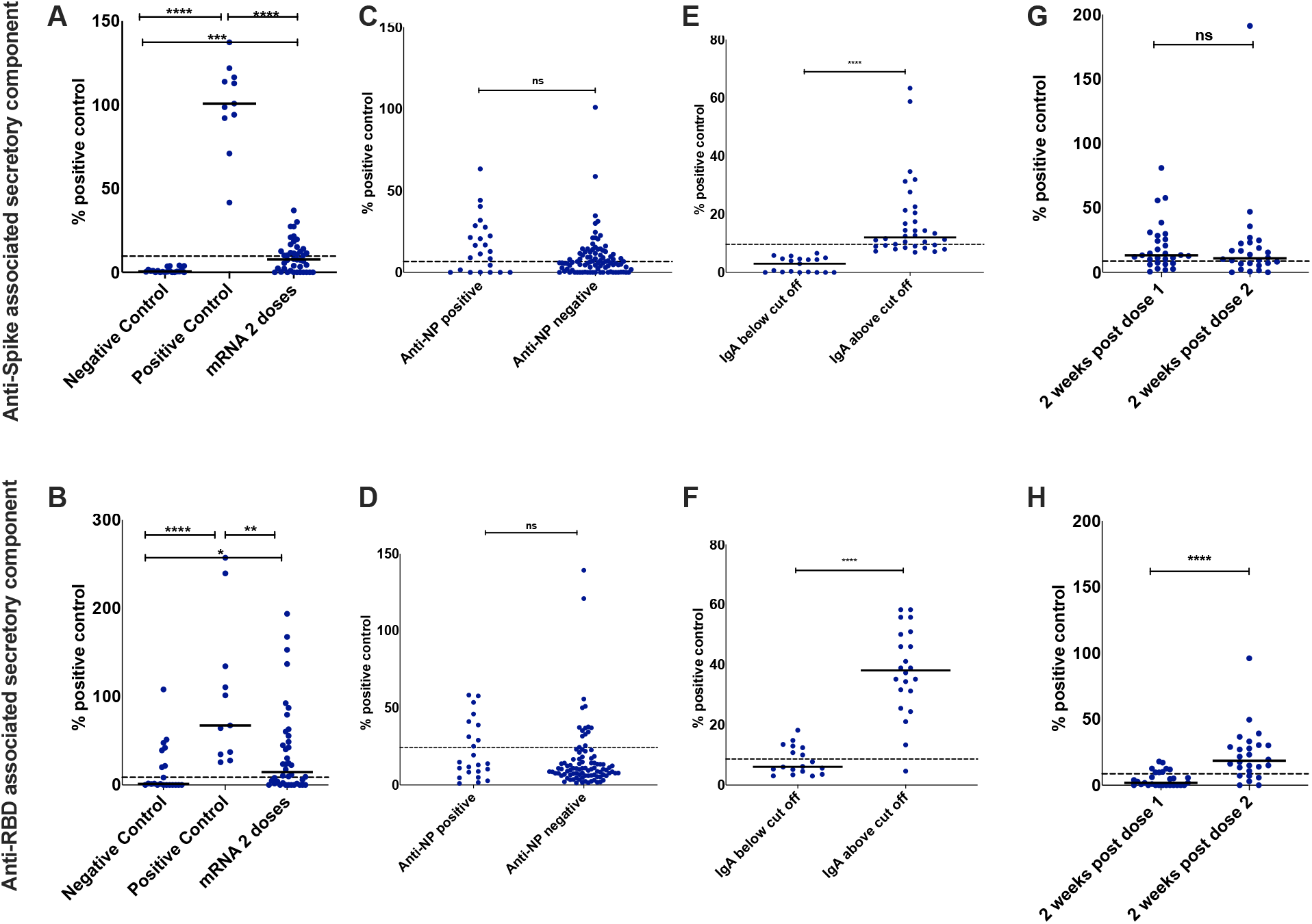
Detection of secretory component associated with anti-Spike and anti-RBD antibodies in saliva from participants receiving COVID-19 mRNA vaccines. An ELISA-based method was used to detect secretory component associated with anti-Spike (**A**) and anti-RBD (**B**) antibodies in the saliva of 2-dose vaccinated subjects (n=93), as well as saliva taken from COVID-19 negative and positive patients (n=77 and 75, respectively). Subjects receiving 2 doses of BNT162b2 or mRNA1273 (**C,D**) were further grouped based on anti-NP antibody status, which is indicative of previous infection. Sub-setting of 2-dose vaccinated subjects into those that were considered above (n=34) or below (n=19) the positive cut off for salivary IgA analyzed for secretory component (**E,F**). Secretory component associated with anti-spike (**G**) and anti-RBD (**H**) antibodies in samples collected post-dose 1 and dose 2 from both MSB-1 (n=27) and MSB-2 (n=36). Solid black bars denote the median for each cohort, while the dotted black line denotes the positive cutoff, calculated as 2 standard deviations above the mean of a pool of negative control samples Mann-Whitney U test was used to calculate significance, with ns=not significant, **=p<0.01, ***=p<0.001 and ****=p<0.0001.

**Fig 4.**
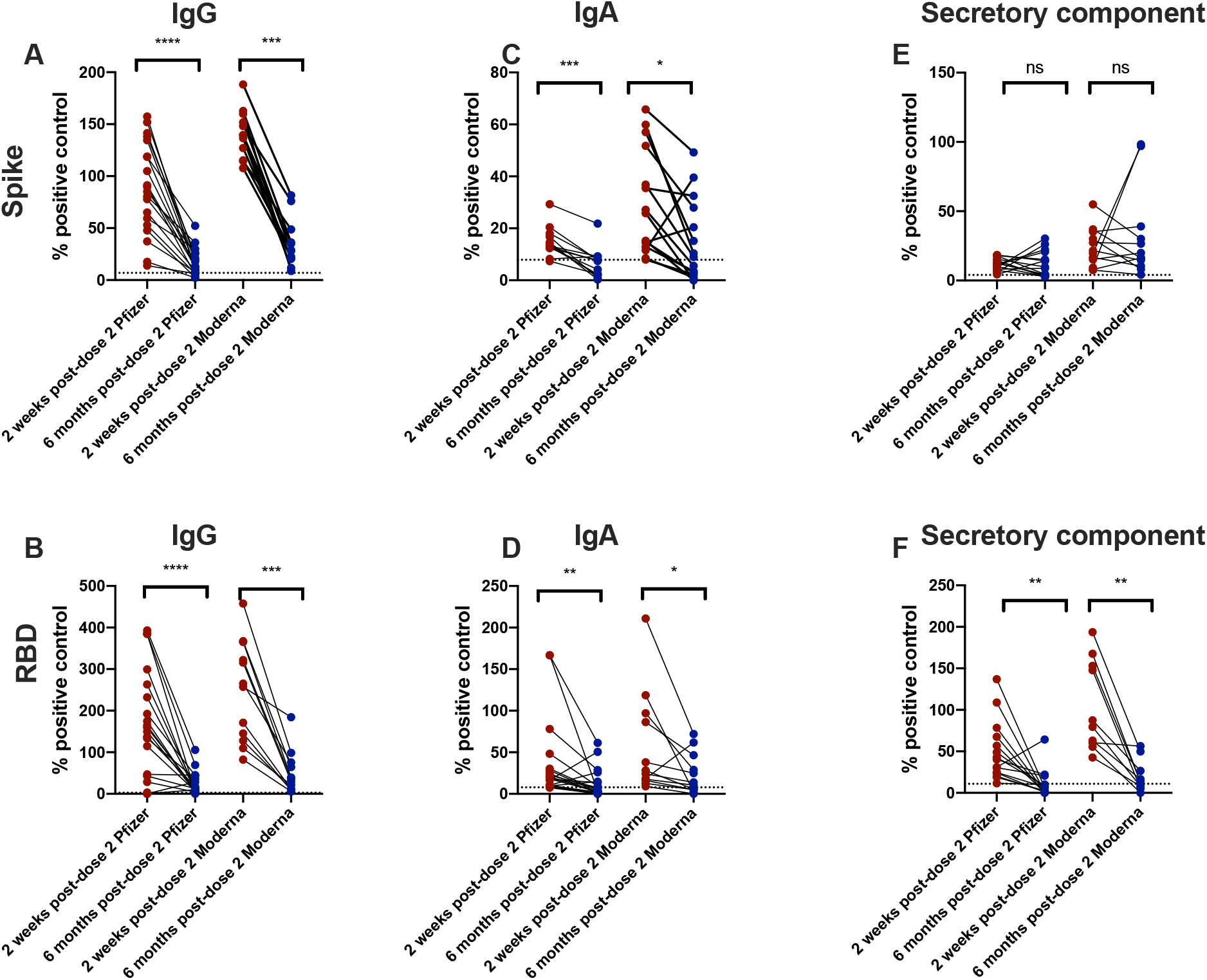
Different decay kinetics of anti-Spike and anti-RBD IgG versus IgA in saliva from participants receiving COVID-19 mRNA vaccines. Saliva from LTCH participants was assessed for the presence of IgG (**A, B**), IgA (**C, D)** and sIgA (**E, F**) antibodies against Spike (A-C) or RBD (D-F). The Wilcoxon signed-rank test was used to calculate significance between groups. ns= not significant, *=p<0.05, **=p<0.01, ***=p<0.001, and ****=p<0.0001.

### Decay kinetics of anti-Spike/RBD IgG and IgA and neutralizing activity in the saliva of COVID-19 mRNA vaccinated participants

Since the LTCH workers were the earliest recipients of mRNA vaccines during the Canadian roll-out, we had the opportunity to examine the stability of anti-Spike/RBD IgG and IgA in the saliva of a subset of the LTCH cohort who are now reaching 6 months post-dose 2. We examined the level of salivary Ab only in those participants who had remained anti-Spike IgA positive at 2-weeks post-dose 2 (n=45, approximately 30% of participants). Using a paired analysis, we observed a significant decline in antigen-specific IgG and IgA levels in the saliva at this time point compared to 2-weeks post-dose 2 (Fig. 4A-D). Although low, anti-Spike Ab associated with the secretory component remained stable in both BNT162b2 and mRNA-1273 vaccinated subjects, although anti-RBD secretory component associated antibodies were significantly reduced at 6 months post-dose 2 (Fig. 4E-F).

In addition, we assessed if COVID-19 mRNA vaccination provokes neutralizing activity against SARS-CoV-2 in the oral cavity, and if this would also be subject to decay. Specifically, saliva at two-fold dilutions was added to hACE2-mCherry expressing HEK293 cells that were co-incubated with recombinant Vesicular Stomatitis Virus (rVSV)-eGFP-SARS-CoV-2-Spike. Infection of HEK293 cells was measured by fluorescence over the course of 72 hours, with prevention of infection by added saliva read out as a green fluorescence reduction in neutralization titer (FRNT). We observed that although saliva from LTCH participants taken at 2-4 weeks post-dose 2 had variable capacity to prevent viral entry into hACE2^+^ HEK293 cells, this was significantly reduced at 6 months post-dose 2 (Supplemental Fig. 5).

Taken together, most Ab against SARS-CoV-2 in the saliva, as well as neutralizing capacity, significantly decline over a 6-month period, except for sIgA against Spike.

### Participants who experience a breakthrough infection have lower levels of vaccination-induced anti-Spike IgA

Correlates of protection against SARS-CoV-2 breakthrough infection are ill-described and have only been examined for IgG^15^. Given that we have detected anti-Spike/RBD IgA in the serum and saliva of mRNA vaccinated participants, we examined whether IgA levels may be associated with protection against breakthrough infection. On 20th April 2021, an outbreak of P.1 lineage SARS-CoV-2 (gamma variant) was declared in a Toronto LTCH home^16^. The residents, who were all doubly vaccinated with mRNA-1273, were part of a separate serum antibody study following LTCH residents that included sampling of blood, but not saliva, at 2-4 weeks post-dose 2 (received on February 2, 2021). Since anti-Spike and anti-RBD IgA levels correlate with each other at 2-4 weeks post-dose 2 (Supplementary Fig. 2), we also measured serum anti-Spike IgA at this time point as a proxy of the salivary IgA response. In the context of this isolated outbreak where n=5 residents were infected we did not see a significant difference between exposed infected vs exposed uninfected participants in terms of levels of anti-Spike and anti-RBD IgG at 2-4 weeks post-dose 2 (Fig. 5A). We noted a trend of reduced anti-Spike and anti-RBD IgA levels in exposed infected vs exposed uninfected participants that reached significance but did not survive a post-hoc multiple test correction (Fig. 5B).

**Fig. 5.**
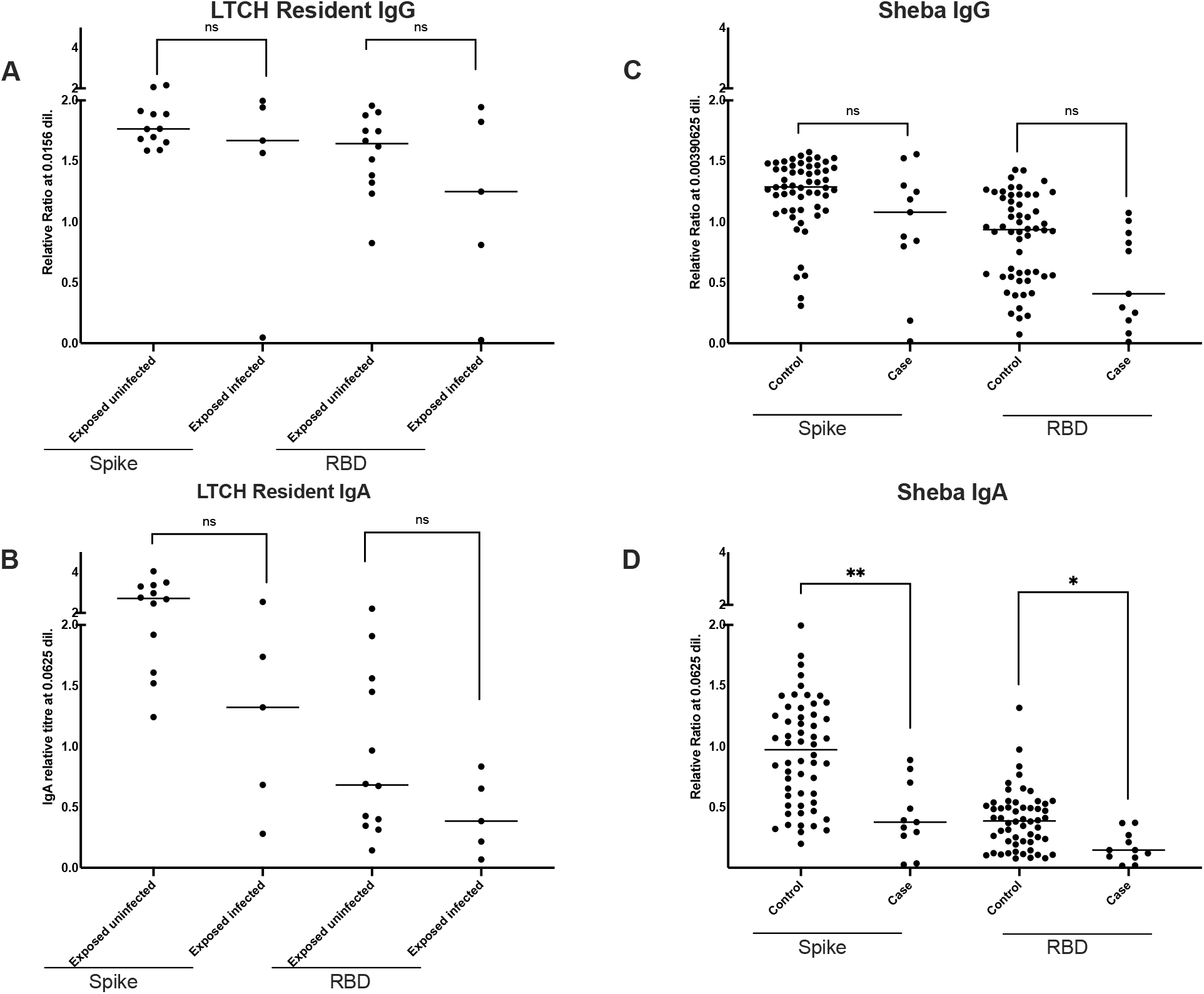
Participants who experience a breakthrough infection have lower level of anti-Spike/RBD IgA at 2-4 weeks post-vaccination: Serum samples from vaccinated Toronto LTCH residents were taken at 2-4 weeks post-dose 2. Serum anti-Spike IgG (**A**) and IgA (**B**) levels were compared in participants who were subsequently exposed to P.1 gamma SARS-CoV-2 and either infected (exposed infected) or not infected (exposed uninfected). In a separate cohort of double vaccinated health care workers from the Sheba Medical Center in Ramat Gan, Israel, serum samples were taken at 2-4 weeks post-dose 2. Serum anti-Spike IgG (**C**) and IgA (**D**) levels were compared in participants who experienced a breakthrough infection (cases) vs controls who did not. Solid black lines denote the median for each cohort. Kruskal-Wallis test with a correction for multiple comparisons was used to conduct statistical analysis between groups. NS=not significant, *=p<0.5, and **=p<0.01.

Given the number of breakthrough infections in this isolated outbreak was small but nevertheless yielded a trend towards reduced anti-Spike/RBD IgA levels at 2-4 weeks post-dose 2 in exposed infected vs exposed uninfected participants, we endeavored to test a replication cohort. We therefore measured serum antibody levels at 2-4 weeks post-dose 2 in a larger case-control cohort of health care workers (HCW) who had received 2 doses of BNT162b2 at the Sheba Medical Center in Ramat Gan, Israel, with the majority of cases infected with the alpha SARS-CoV-2 variant^15^. We found that anti-Spike/RBD IgG levels were modestly but not significantly lower in the serum of cases versus controls (Fig. 5C). In contrast, anti-Spike and anti-RBD IgA levels were both significantly lower in cases vs controls, and this held up to a multiple correction using a Kruskal Wallace Dunn’s multiple comparison test (Fig. 5D). Thus, despite different vaccine regimes (mRNA-1273 vs BNT162b2), geographical locations (Canada vs Israel) and viral exposures (gamma vs alpha), in both cohorts anti-Spike/RBD serum IgA but not IgG levels are lower at 2-4 weeks post-dose 2 in participants who subsequently are infected with SARS-CoV-2.

## Discussion

In this study we observed robust IgG levels to Spike/RBD in the saliva of participants immunized with BNT162b2 or mRNA-1273 that correlate with the systemic IgG response, and these IgG are significantly diminished at 6 months post-dose 2. A different picture emerges, however, for salivary anti-Spike/RBD IgA. In nearly all participants we observed a modest sIgA response to Spike/RBD in the saliva after a single dose of mRNA vaccine which is maintained in only approximately 30% of participants after dose 2. Moreover, in these subjects, although the sIgA response to Spike is modest, it is more resilient to decay. This preserved sIgA response may be very important for preventing breakthrough infections. Indeed, vaccinated participants who subsequently experience a SARS-CoV-2 infection have significantly lower levels of anti-Spike serum IgA at 2-4 weeks post-dose 2 compared to subjects who remain uninfected.

Our findings of IgA in the saliva are consistent with recent reports showing that after two doses of BNT162b2 or mRNA-1273 anti-Spike and anti-RBD Ab could be detected in saliva^17-19^. We also distinguish between locally produced and systemically derived IgA using secretory component detection. It is unclear how anti-Spike/RBD sIgA is generated in the saliva following i.m. immunization. Of note, Spike protein can be detected in the plasma, increasing one to 5 days after mRNA-1273 vaccination using an ultra-sensitive detection technique^20^. Thus one possibility is that plasma-associated Spike antigen may reach the salivary glands (which are surrounded by capillaries^21^), provoking a local sIgA response. Another possibility is that a mucosal IgA response to mRNA vaccination takes place in the gut, and plasma cells generated at this location leave the gut (as we have shown before^22^), disseminating to other mucosal surfaces such as the oral cavity. Indeed, expression of the gut homing integrin α4β7 on circulating immune cells has been observed following administration of yellow fever^23^ and cholera toxin vaccines^24^ via the systemic route in humans. Recirculation between mucosal compartments could explain the sustained levels of anti-Spike sIgA in the saliva at 6 months post-boost which is not observed for the systemic IgG response. Animal models are ideal for obtaining further insights into mechanisms that explain how SARS-CoV-2 mRNA vaccines provoke antigen-specific sIgA production at mucosal surfaces.

There are some limitations to our study: we did not have saliva available from the LTCH residents nor the Sheba HCW for confirmation that the association of high serum anti-Spike IgA levels with protection against breakthrough infection was also observed in a mucosal secretion. Finding a prospective cohort where saliva was collected at 2-4 weeks post-dose 2 where breakthrough infections are monitored, a rarity in and of itself, is now confounded by circulation of Omicron which is highly immune evasive. However, we observed that the levels of anti-Spike and anti-RBD IgA in the saliva at 2 weeks post-dose 2 correlates with IgA levels in the serum (Supplementary Fig. 2), suggesting that what we are detecting in the serum at least partially mirrors the oral compartment. Another limitation of our study is that we do not yet have 6-month samples examining the level of anti-Spike/RBD antibodies in the saliva of subjects who had a longer interval between dose 1 and dose 2. It will be of interest to determine if dose interval impacts salivary antibody decay rates. Lastly, it is unclear whether anti-Spike sIgA antibodies in the saliva will further erode after 6 months post-dose 2 or will remain persistent.

Dimeric secretory IgA has been shown to have potent neutralization activity against SARS-CoV-2 *in vitro*^*6*^. Using a flow cytometry-based pseudovirus entry assay we found that saliva from COVID-19 mRNA vaccinated subjects can variably prevent pseudovirus infection of ACE2^+^ cells. This is consistent with work by Nahass et al who have examined neutralizing capacity of saliva in response to different vaccine regimes^25^. However, aside from direct neutralization, other IgA-dependent mechanisms may be relevant to protection against breakthrough infection. For example, IgA may serve to “enchain” microbes as it does in other mucosal sites such as the gut^9^, and even non-mucosal sites such as the brain meninges^26^. Indeed, although the neutralization activity of saliva at 6 months post-dose 2 was greatly diminished, anti-Spike sIgA levels had not significantly decayed at this time-point. It will be of interest to examine structural interactions between sIgA dimers with SARS-CoV-2 in the context of the mucosae.

In summary, we provide evidence that anti-Spike sIgA are induced and maintained in the saliva of approximately 30% of mRNA vaccinated participants and that high anti-Spike/RBD serum IgA levels are associated with protection against subsequent breakthrough infection. Intranasal (i.n.) adenoviral-based vaccines that robustly induce anti-Spike/RBD IgA have been shown to prevent transmission in golden hamsters^27^. Moreover, a SARS-CoV-2 mRNA vaccine i.m. prime followed by an adenoviral i.n. boost provides superior protection against infection compared to i.m. only regimes, even against divergent variants such as B.1.1.351 (beta)^28^. An i.m. prime followed by i.n. boost strategy is worth considering for preventing person-to-person transmission of SARS-CoV-2 and for broad protection against emerging variants.

## Data Availability

The data will be available upon reasonable request.

## Acknowledgments

Dr. Yves Durocher at the National Research Council of Canada (NRC) kindly donated reagents for the serum assays. We thank Elizabeth Yue, Antonio Estacio, Serena Loklam Chau, Ryan Law and Eric Yixiao Cao for LTCH sample processing and sustaining the infrastructure of sample processing in the Ostrowski lab. We thank Jeff Browning for critical reading of this manuscript. We would like to thank Florian Krammer for advice on secretory component detection by ELISA. Funding for this study was from a Foundation grant from the Canadian Institutes of Health Research (JG, Fund #15992), a COVID-19 Immunity Task force grant (SS, AM, MO, ACG and JG), an “Ontario Together” province of Ontario grant (JG and ACG), a Donation from the Royal Bank of Canada (RBC) Donation from the Krembil Foundation to the Sinai Health System Foundation.

