## Supplemental Figures and Tables for "Systemic and mucosal IgA responses are variably induced in response to SARS-CoV-2 mRNA vaccination and are associated with protection against subsequent infection"

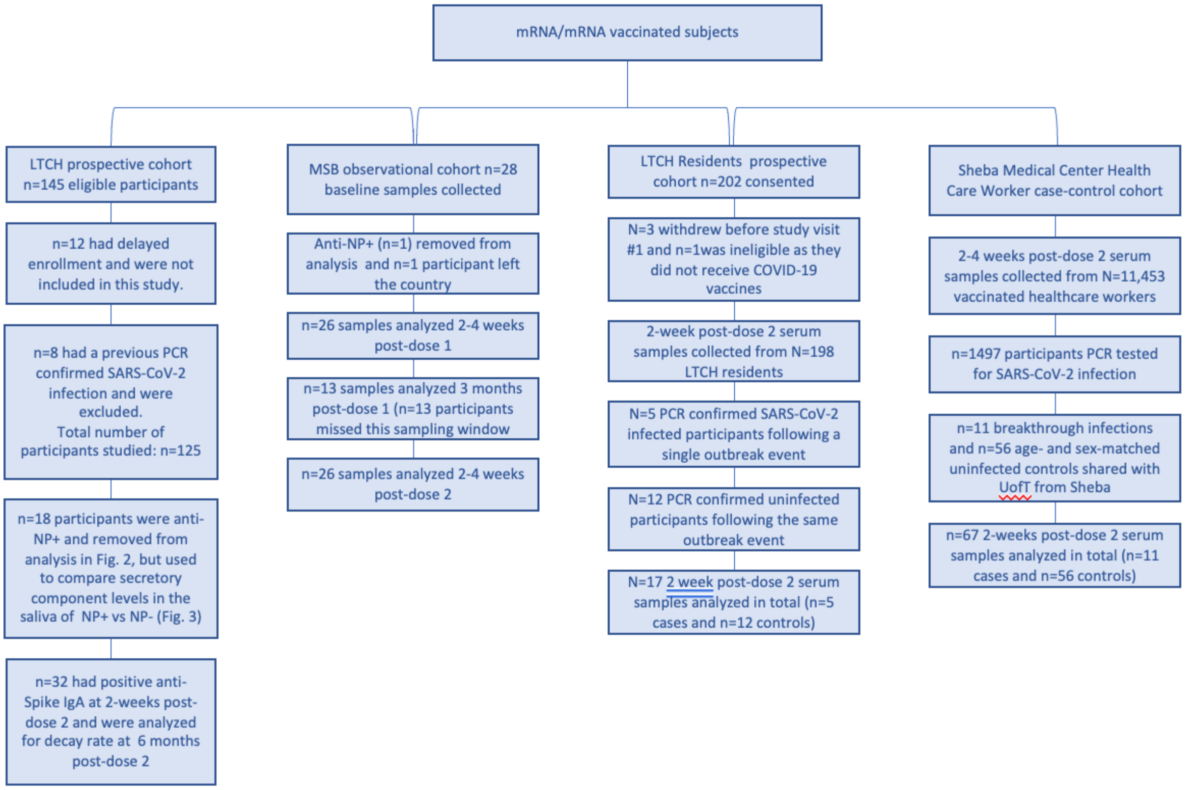


**Supplementary Fig. 1. Study Design**

Two observational cohorts (LTCH staff, MSB) a prospective cohort (LTCH Residents), and a case-control cohort (Sheba Medical Center HCW) were integrated into our overall study of mRNA/mRNA vaccinated subjects.

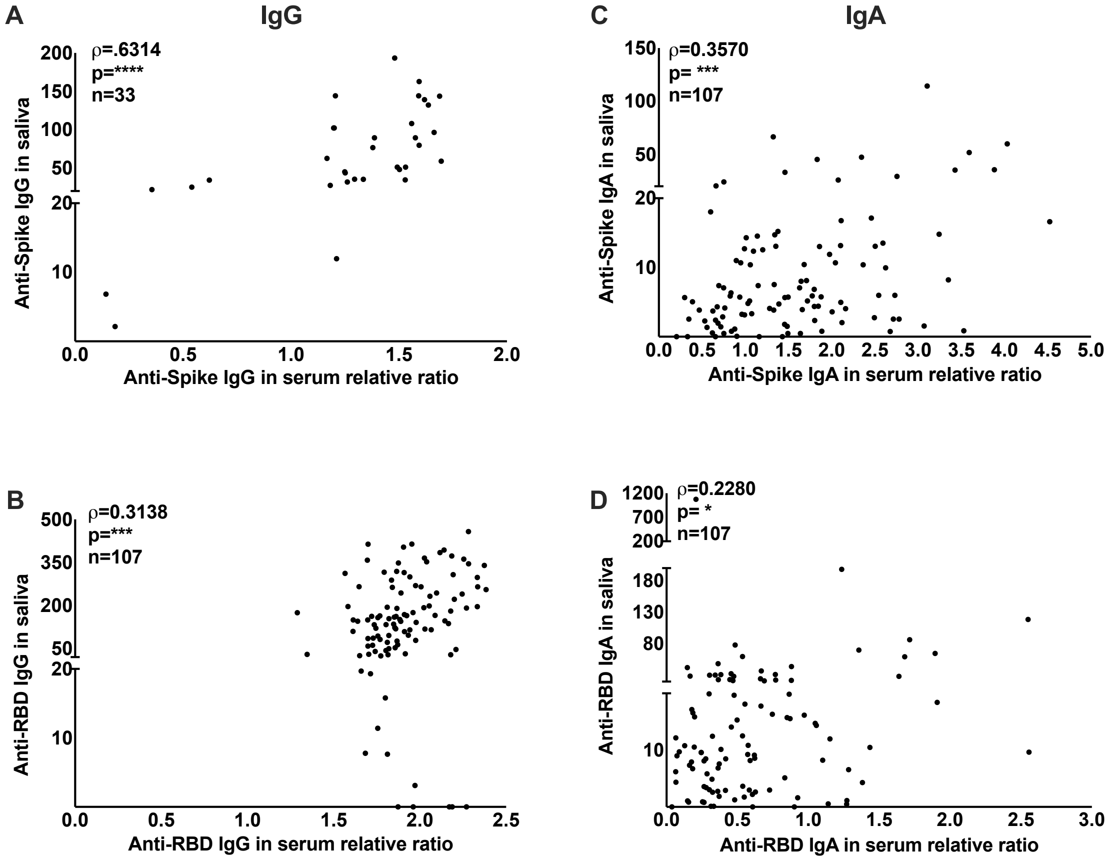


**Supplementary Fig. 2. Correlation of anti-spike and anti-RBD antibodies in the saliva and serum 2-4 weeks after mRNA vaccination**

Subjects from the LTCH cohort were sampled 2-4 weeks after their second dose of COVID-19 mRNA vaccination. Anti-Spike (**A,C**) and anti-RBD (**B,D**) IgG and IgA were analyzed in serum and saliva via an ELISA-based method. A Spearman test was used to calculate the correlation coefficient for each plot. During analysis of anti-Spike IgG (**A**) in the serum of LTCH subjects, we observed oversaturation of signal, and thus repeated the analysis for anti-Spike IgG using a lower concentration on a subset (n=33) of samples which is what is shown here. *=p<0.05, ***=p<0.001 and ****=p<0.0001.

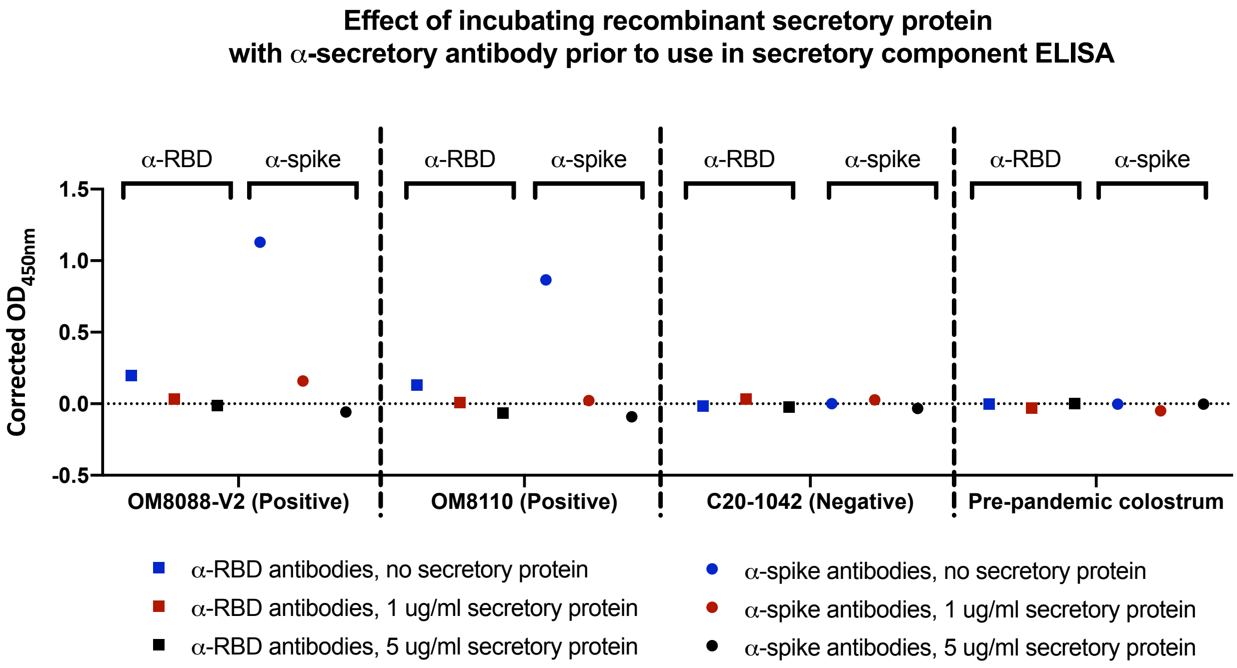


**Supplementary Fig. 3. Detection of secretory component associated with Spike/RBD-specific antibodies from COVID-19 patient saliva**

Validation of secretory component assay was confirmed by outcompeting signal with recombinant secretory component. We observed that incubation of COVID-19 positive samples with recombinant secretory component prior to incubation in the experimental plate brought signal down to negative control levels in a concentration-dependent manner. Pre-pandemic colostrum showed no evidence of anti-Spike/RBD associated secretory component signal, as expected.

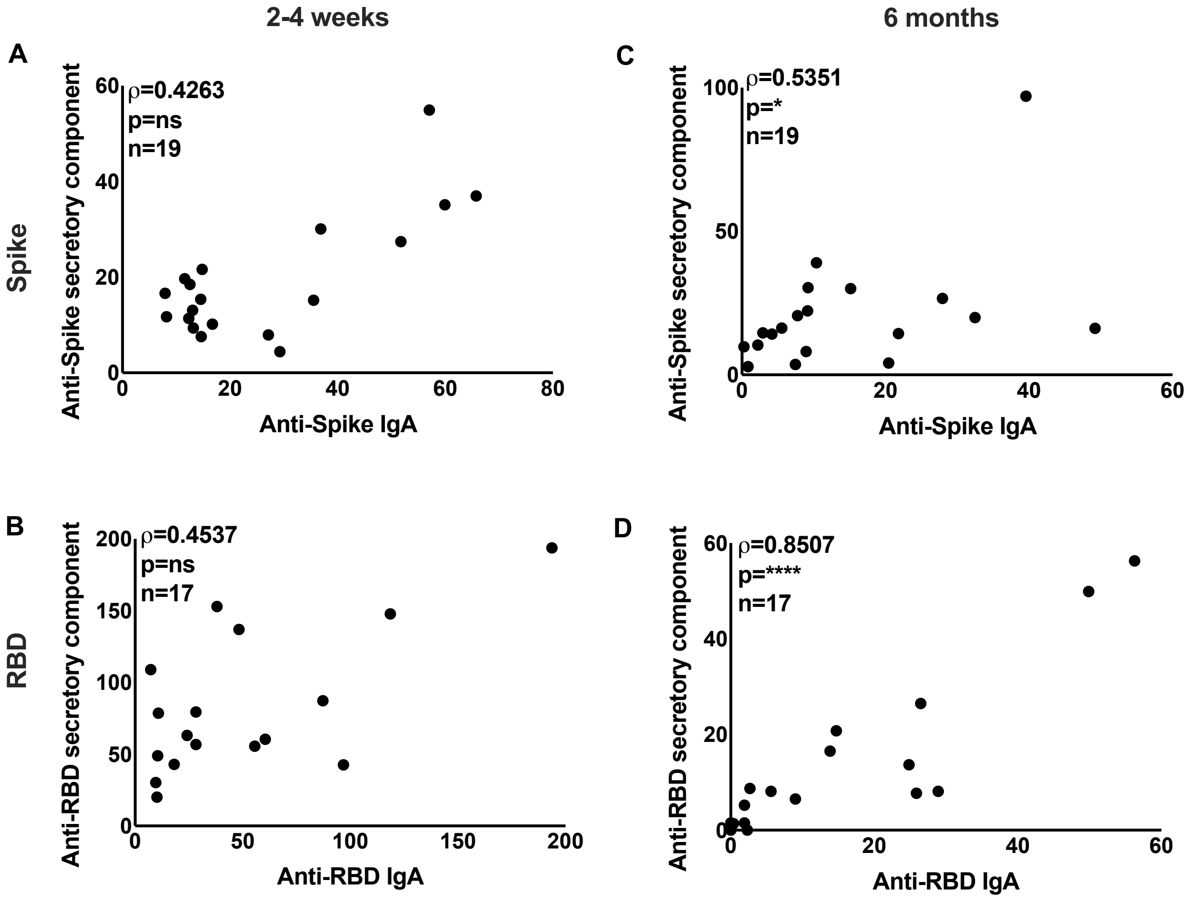


**Supplementary Fig. 4. Correlation of anti-spike and anti-RBD IgA and secretory component in the saliva at 2 weeks and 6 months post-dose 2.**

Participants from the LTCH cohort were sampled 2-4 weeks (A,B) and 6 months (C,D) after their second dose of COVID-19 mRNA vaccination. IgA and secretory component were analyzed in saliva via an ELISA-based method. A Spearman test was used to calculate the correlation coefficient for each plot, examining subjects who were positive for anti-Spike IgA at 2 weeks post-dose 2. ns= not significant, *=p<0.05 and ****=p<0.0001.

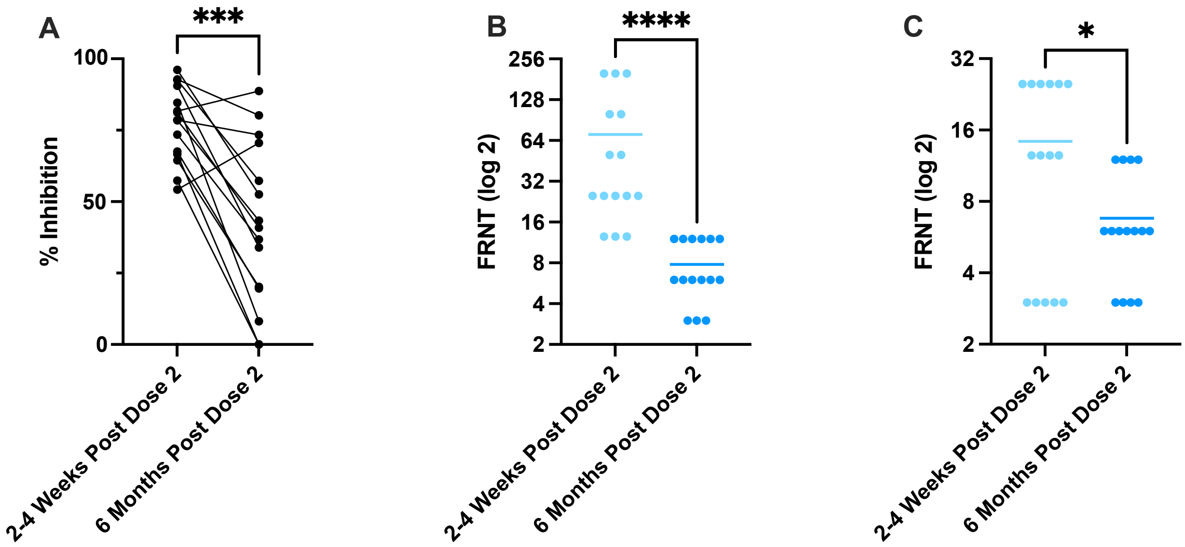


**Supplementary Fig 5. Neutralizing antibodies detected in the saliva of participants receiving COVID-19 mRNA vaccines.**
Saliva taken from a subset (n=15) of mRNA-vaccinated subjects representing a range of IgA titers 2-4 weeks post-dose 2 was assessed for the presence of neutralizing antibodies and results were expressed as a percentage of maximum infection (**A),** where a lower percentage indicates decreased infection of hACE-2 HEK293 cells by rVSV-eGFP-SARS-CoV-2-Spike. Fluorescent reduction neutralization titre (FRNT) using a cut off of either 50% (**B**) or 70% (**C**) from n=15 participants at 2-4 weeks and 6 months post-dose 2 was also analyzed. The Wilcoxon signed-rank test was used to calculate significance between groups, where *=p<0.05, ***=p<0.001, and ****=p<0.0001.

**Supplementary Table 1.**
LTCH cohort (mRNA/mRNA)

| Participant ID | Age | Sex | Vaccine type | Interval between dose 1 and dose 2 (weeks) |
|---|---|---|---|---|
| 1 | 46-50 | F | mRNA1273 | 4 |
| 2 | 61-65 | F | mRNA1273 | 4.7 |
| 3 | 46-50 | F | BNT162b2 | 5.9 |
| 4 | 41-45 | F | mRNA1273 | 4 |
| 5 | 56-60 | F | BNT162b2 | 3 |
| 6 | 41-45 | F | mRNA1273 | 4 |
| 7 | 56-60 | F | mRNA1273 | 4 |
| 8 | 41-45 | F | BNT162b2 | 5 |
| 9 | 56-60 | F | mRNA1273 | 4 |
| 10 | 46-50 | F | mRNA1273 | 4 |
| 11 | 36-40 | F | BNT162b2 | 5 |
| 12 | 51-55 | M | BNT162b2 | 2.9 |
| 13 | 56-60 | F | mRNA1273 | 2.9 |
| 14 | 46-50 | F | BNT162b2 | 5.6 |
| 15 | 31-35 | F | BNT162b2 | 5.3 |
| 16 | 51-55 | F | BNT162b2 | 5.6 |
| 17 | 61-65 | F | BNT162b2 | 47.7 |
| 18 | 51-55 | F | BNT162b2 | 4.4 |
| 19 | 51-55 | F | mRNA1273 | 4 |
| 20 | 61-65 | F | BNT162b2 | 5 |
| 21 | 56-60 | F | BNT162b2 | 5 |
| 22 | 26-30 | F | BNT162b2 | 5.6 |
| 23 | 56-60 | F | BNT162b2 | 5.6 |
| 24 | 46-50 | F | BNT162b2 | 5.6 |
| 25 | 41-45 | F | BNT162b2 | 5.6 |
| 26 | 36-40 | F | BNT162b2 | 5.6 |
| 27 | 61-65 | F | BNT162b2 | 5.6 |
| 28 | 51-55 | F | BNT162b2 | 5.6 |
| 29 | 41-45 | F | BNT162b2 | 5.6 |
| 30 | 51-55 | M | BNT162b2 | 7 |
| 31 | 61-65 | F | BNT162b2 | 5.6 |
| 32 | 46-50 | F | BNT162b2 | 5 |
| 33 | 26-30 | F | BNT162b2 | 5.6 |
| 34 | 31-35 | M | BNT162b2 | 5.6 |
| 35 | 36-40 | F | mRNA1273 | 4 |
| 36 | 31-35 | F | BNT162b2 | 5 |
| 37 | 41-45 | F | BNT162b2 | 5.6 |

| | | | | |
|---|---|---|---|---|
| 38 | 26-30 | F | BNT162b2 | 5.6 |
| 39 | 41-45 | F | BNT162b2 | 5 |
| 40 | 61-65 | F | BNT162b2 | 6.4 |
| 41 | 36-40 | M | BNT162b2 | 5.6 |
| 42 | 31-35 | F | mRNA1273 | 4 |
| 43 | 36-40 | M | mRNA1273 | 4 |
| 44 | 51-55 | F | mRNA1273 | 3 |
| 45 | 46-50 | M | mRNA1273 | 4 |
| 46 | 26-3- | F | mRNA1273 | 4 |
| 47 | 56-60 | F | mRNA1273 | 4 |
| 48 | 31-35 | F | mRNA1273 | 5 |
| 49 | 56-60 | M | mRNA1273 | 4.9 |
| 50 | 51-55 | F | mRNA1273 | 5 |
| 51 | 41-45 | F | mRNA1273 | 4.1 |
| 52 | 51-55 | F | mRNA1273 | 2.9 |
| 53 | 36-40 | F | BNT162b2 | 4.6 |
| 54 | 41-45 | F | BNT162b2 | 4.1 |
| 55 | 61-65 | F | BNT162b2 | 3.4 |
| 56 | 61-65 | F | BNT162b2 | 4.6 |
| 57 | 20-25 | | BNT162b2 | 3.7 |
| 58 | 20-25 | F | BNT162b2 | 4.9 |
| 59 | 31-35 | F | BNT162b2 | 3 |
| 60 | 26-30 | F | BNT162b2 | 4.4 |
| 61 | 31-35 | F | BNT162b2 | 3.7 |
| 62 | 46-50 | F | BNT162b2 | 4.9 |
| 63 | 36-40 | M | BNT162b2 | 3 |
| 64 | 61-65 | M | BNT162b2 | 4.6 |
| 65 | 46-50 | F | BNT162b2 | 3.1 |
| 66 | 61-65 | F | BNT162b2 | 5 |
| 67 | 26-30 | F | BNT162b2 | 4.7 |
| 68 | 51-55 | F | BNT162b2 | 4.4 |
| 69 | 46-50 | F | BNT162b2 | 3 |
| 70 | 41-45 | F | BNT162b2 | 3.9 |
| 71 | 51-55 | F | BNT162b2 | 3.9 |
| 72 | 31-35 | F | BNT162b2 | 8.4 |
| 73 | 46-50 | F | BNT162b2 | 6.9 |
| 74 | 51-55 | F | BNT162b2 | 3 |
| 75 | 20-25 | F | BNT162b2 | 3.1 |
| 76 | 20-25 | | BNT162b2 | 3.7 |
| 77 | 51-55 | F | BNT162b2 | 3 |
| 78 | 56-60 | F | BNT162b2 | 4.7 |
| 79 | 20-25 | F | BNT162b2 | 3.1 |
| 80 | 55-60 | F | BNT162b2 | 5.1 |

| | | | | |
|---|---|---|---|---|
| 81 | 36-40 | M | BNT162b2 | 5 |
| 82 | 26-30 | F | BNT162b2 | 5 |
| 83 | 56-60 | F | BNT162b2 | 4 |
| 84 | 20-25 | F | mRNA1273 | 4 |
| 85 | 20-25 | F | BNT162b2 | 5 |
| 86 | 46-50 | M | mRNA1273 | 3.4 |
| 87 | 41-45 | F | BNT162b2 | 5 |
| 88 | 66-70 | F | BNT162b2 | 5 |
| 89 | 36-40 | M | mRNA1273 | 3 |
| 90 | 31-35 | F | BNT162b2 | 3 |
| 91 | 31-35 | F | mRNA1273 | 3.4 |
| 92 | 20-25 | F | BNT162b2 | 3 |
| 93 | 26-30 | F | mRNA1273 | 3 |
| 94 | 20-25 | F | BNT162b2 | 3 |
| 95 | 41-45 | F | BNT162b2 | 3 |
| 96 | 61-65 | F | mRNA1273 | 3 |
| 97 | 56-60 | F | mRNA1273 | 4 |
| 98 | 41-45 | F | BNT162b2 | 49.1 |
| 99 | 26-30 | F | BNT162b2 | 49.1 |
| 100 | 56-60 | F | mRNA1273 | 3 |
| 101 | 26-30 | F | mRNA1273 | 4 |
| 102 | 36-40 | F | BNT162b2 | 3 |
| 103 | 61-65 | F | mRNA1273 | 3.6 |
| 104 | 36-40 | F | BNT162b2 | 5 |
| 105 | 26-30 | F | mRNA1273 | 3.4 |
| 106 | 20-25 | F | mRNA1273 | 4 |
| 107 | 71-75 | F | mRNA1273 | |
| **Summary** | **Median=46** | **%M=11**<br>**%F=89** | **%mRNA1273=32**<br>**%BNT162b2=68** | **Median=4.4** |

**Supplementary Table 2a.**
COVID-19 era negative controls

| Participant ID | Age | Sex | Collection period | Used in Fig 1, Fig. 3 or as pooled negative |
|---|---|---|---|---|
| 108 | 26-30 | M | COVID era | Fig. 3 |
| 109 | 20-25 | F | COVID era | Fig. 3 |
| 110 | 61-65 | M | COVID era | Fig. 3 |
| 111 | 36-40 | M | COVID era | Fig. 3 |
| 112 | 56-6- | F | COVID era | Fig. 3 |
| 113 | 41-45 | M | COVID era | Fig. 3 |
| 114 | 46-50 | F | COVID era | Fig. 3 |
| 115 | 26-30 | M | COVID era | Fig. 3 |
| 116 | 31-35 | F | COVID era | Fig. 3 |
| 117 | 31-35 | M | COVID era | Fig. 3 |
| 118 | 36-40 | F | COVID era | Fig. 3 |
| 119 | 36-40 | M | COVID era | Fig. 3 |
| 120 | 31-35 | M | COVID era | Fig. 3 |
| 121 | 36-40 | M | COVID era | Fig. 3 |
| 122 | 66-70 | F | COVID era | Fig. 3 |
| 123 | 66-70 | M | COVID era | Fig. 3 |
| 124 | 91-95 | F | COVID era | Fig. 3 |
| 125 | 36-40 | M | COVID era | Fig. 3 |
| 126 | 71-75 | F | COVID era | Fig. 3 |
| 127 | 66-70 | M | COVID era | Fig. 3 |
| 128 | 36-40 | F | COVID era | Fig. 3 |
| 129 | 41-45 | M | COVID era | Fig. 3 |
| 130 | 41-45 | M | COVID era | Fig. 3 |
| 131 | 51-55 | M | COVID era | Fig. 3 |
| 132 | 61-65 | M | COVID era | Fig. 3 |
| 133 | 56-60 | F | COVID era | Fig. 3 |
| 134 | 61-65 | M | COVID era | Fig. 3 |
| 135 | 71-75 | M | COVID era | Fig. 3 |
| 136 | 81-85 | F | COVID era | Fig. 3 |
| 137 | 51-55 | M | COVID era | Fig. 3 |
| 138 | 71-75 | M | COVID era | Fig. 3 |
| 139 | 61-65 | F | COVID era | Fig. 3 |
| 140 | 61-65 | M | COVID era | Fig. 3 |
| 141 | 81-85 | F | COVID era | Fig. 3 |
| 142 | 46-50 | M | COVID era | Fig. 3 |
| 143 | 66-70 | F | COVID era | Fig. 3 |
| 144 | 20-25 | F | COVID era | Fig. 3 |
| 145 | 66-70 | F | COVID era | Fig. 3 |

| | | | | |
|---|---|---|---|---|
| 146 | 26-30 | M | COVID era | Fig. 3 |
| 147 | 61-65 | F | COVID era | Fig. 3 |
| 148 | 61-65 | M | COVID era | Fig. 3 |
| 149 | 41-45 | M | COVID era | Fig. 3 |
| 150 | 61-65 | M | COVID era | Fig. 3 |
| 151 | 86-90 | M | COVID era | Fig. 3 |
| 152 | 81-85 | F | COVID era | Fig. 3 |
| 153 | 81-85 | M | COVID era | Fig. 3 |
| 154 | 76-80 | M | COVID era | Fig. 3 |
| 155 | 56-60 | M | COVID era | Fig. 3 |
| 156 | 56-60 | M | COVID era | Fig. 3 |
| **Summary** | **Median age=58** | **%M=63 %F=37** | | |

**Supplementary Table 2b.**
Pre-COVID era negative controls

| Sample ID | Age | Sex | Used in Fig. 1, Fig. 3 or as pooled negative |
|---|---|---|---|
| 157 | 31-35 | F | Pooled negative |
| 158 | 41-45 | F | Pooled negative |
| 159 | 41-45 | M | Pooled negative |
| 160 | 31-35 | F | Pooled negative |
| 161 | 66-70 | M | Pooled negative |
| 162 | 20-25 | F | Pooled negative |
| 163 | 31-35 | F | Pooled negative |
| 164 | 26-30 | F | Pooled negative |
| 165 | 41-45 | F | Pooled negative |
| 166 | 31-35 | M | Pooled negative |
| 167 | 26-30 | M | Pooled negative |
| 168 | 41-45 | F | Pooled negative |
| 169 | 46-50 | F | Pooled negative |
| 170 | 36-40 | F | Pooled negative |
| 171 | 56-60 | M | Pooled negative |
| 172 | 36-40 | M | Pooled negative |
| 173 | 51-55 | F | Pooled negative |
| 174 | 15-19 | M | Pooled negative |
| 175 | 20-25 | M | Pooled negative |
| 176 | 46-50 | M | Pooled negative |
| 177 | 36-40 | F | Pooled negative |
| 178 | 36-40 | F | Pooled negative |
| 179 | 46-50 | F | Pooled negative |
| 180 | 41-45 | F | Pooled negative |
| 181 | 46-50 | M | Pooled negative |
| 182 | 51-55 | F | Pooled negative |
| 183 | 31-35 | M | Pooled negative |
| 184 | 31-35 | F | Pooled negative |
| 185 | 56-60 | F | Pooled negative |
| 186 | 51-55 | F | Pooled negative |
| 187 | 26-30 | F | Pooled negative |
| 188 | 36-40 | M | Pooled negative |
| 189 | 41-45 | M | Pooled negative |
| 190 | 31-35 | F | Pooled negative |
| 191 | 26-30 | F | Pooled negative |
| 192 | 36-40 | F | Pooled negative |
| 193 | 56-60 | M | Pooled negative |
| 194 | 41-45 | M | Pooled negative |
| 195 | 41-45 | F | Pooled negative |
| 196 | 36-40 | F | Pooled negative |
| 197 | 31-35 | M | Pooled negative |

| | | | |
|---|---|---|---|
| 198 | 36-40 | M | Pooled negative |
| 199 | 46-50 | F | Pooled negative |
| 200 | 41-45 | F | Pooled negative |
| 201 | 15-19 | F | Pooled negative |
| 202 | 26-30 | F | Pooled negative |
| 203 | 21-25 | M | Pooled negative |
| 204 | 36-40 | F | Pooled negative |
| 205 | 36-40 | M | Pooled negative |
| 206 | 31-35 | F | Pooled negative |
| 207 | 26-30 | F | Pooled negative |
| 208 | 31-35 | M | Fig. 3 + pooled negative |
| 209 | 32-35 | M | Fig. 3 + pooled negative |
| 210 | 41-45 | M | Fig. 3 + pooled negative |
| 211 | 51-55 | F | Fig. 3 + pooled negative |
| 212 | 56-60 | M | Fig. 3 + pooled negative |
| 213 | 56-60 | F | Fig. 3 + pooled negative |
| 214 | 46-50 | F | Fig. 3 + pooled negative |
| 215 | 41-45 | M | Fig. 3 + pooled negative |
| 216 | 41-45 | M | Fig. 3 + pooled negative |
| 217 | 36-40 | F | Fig. 3 + pooled negative |
| 218 | 46-50 | M | Fig. 3 + pooled negative |
| 219 | 41-45 | F | Fig. 3 + pooled negative |
| 220 | 51-55 | M | Fig. 3 + pooled negative |
| 221 | 41-45 | F | Fig. 3 + pooled negative |
| 222 | 26-30 | M | Fig. 3 + pooled negative |
| 223 | 41-45 | F | Fig. 3 + pooled negative |
| 224 | 26-30 | M | Fig. 3 + pooled negative |
| 225 | 31-35 | M | Fig. 3 + pooled negative |
| 226 | 36-40 | M | Fig. 3 + pooled negative |
| 227 | 41-45 | F | Fig. 3 + pooled negative |
| 228 | 36-40 | F | Fig. 3 + pooled negative |
| 229 | 15-19 | M | Fig. 3 + pooled negative |
| 230 | 20-25 | F | Fig. 3 + pooled negative |
| 231 | 46-50 | F | Fig. 3 + pooled negative |
| 232 | 56-60 | F | Fig. 3 + pooled negative |
| 233 | 61-65 | M | Fig. 3 + pooled negative |
| 234 | 36-40 | F | Fig. 3 + pooled negative |
| **Summary** | **Median age=39** | **%M=42**<br>**%F=58** | |

**Supplementary Table 2c.**
COVID-19 patient data

| Participant ID | Age | Sex | Days PSO | Status | Used in Fig.1 or Fig. 3 |
|---|---|---|---|---|---|
| 235 | 71-75 | M | 46 | Convalescent | Fig. 3 |
| 236 | 31-35 | M | 78 | Convalescent | Fig. 3 |
| 237 | 56-60 | M | 47 | Convalescent | Fig. 3 |
| 238 | 71-75 | F | 42 | Convalescent | Fig. 3 |
| 239 | 26-30 | M | 50 | Convalescent | Fig. 3 |
| 240 | 51-55 | M | 48 | Convalescent | Fig. 3 |
| 241 | 46-50 | M | 60 | Convalescent | Fig. 3 |
| 242 | 41-45 | M | 44 | Convalescent | Fig. 3 |
| 243 | 56-60 | F | 45 | Convalescent | Fig. 3 |
| 244 | 56-60 | F | 39 | Convalescent | Fig. 3 |
| 245 | 51-55 | M | 65 | Convalescent | Fig. 3 |
| 246 | 41-45 | F | 53 | Convalescent | Fig. 3 |
| 247 | 61-65 | F | 51 | Convalescent | Fig. 3 |
| 248 | 56-60 | F | 47 | Convalescent | Fig. 3 |
| 249 | 26-30 | M | 42 | Convalescent | Fig. 3 |
| 250 | 41-45 | M | 49 | Convalescent | Fig. 3 |
| 251 | 56-60 | M | 39 | Convalescent | Fig. 3 |
| 252 | 56-60 | F | 59 | Convalescent | Fig. 3 |
| 253 | 76-80 | M | 64 | Convalescent | Fig. 3 |
| 254 | 51-55 | M | 58 | Convalescent | Fig. 3 |
| 255 | 71-75 | M | 57 | Convalescent | Fig. 3 |
| 256 | 56-60 | F | 58 | Convalescent | Fig. 3 |
| 257 | 61-65 | M | 40 | Convalescent | Fig. 3 |
| 258 | 51-55 | M | 64 | Convalescent | Fig. 3 |
| 259 | 26-30 | F | 91 | Convalescent | Fig. 3 |
| 260 | 51-55 | M | 49 | Convalescent | Fig. 3 |
| 261 | 66-70 | F | 65 | Convalescent | Fig. 3 |
| 262 | 51-55 | F | 52 | Convalescent | Fig. 3 |
| 263 | 51-55 | M | 82 | Convalescent | Both |
| 264 | 51-55 | M | 75 | Convalescent | Fig. 3 |
| 265 | 26-30 | F | 56 | Convalescent | Fig. 3 |
| 266 | 61-65 | M | 31 | Convalescent | Fig. 3 |
| 267 | 76-80 | F | 47 | Convalescent | Fig. 3 |
| 268 | 66-70 | F | 40 | Convalescent | Fig. 3 |
| 269 | 46-50 | M | 50 | Convalescent | Fig. 3 |
| 270 | 81-85 | F | 67 | Convalescent | Fig. 3 |
| 271 | 61-65 | M | 62 | Convalescent | Fig. 3 |
| 272 | 71-75 | M | 80 | Convalescent | Fig. 3 |
| 273 | 56-60 | M | 104 | Convalescent | Fig. 3 |
| 274 | 71-75 | M | 40 | Convalescent | Fig. 3 |

| | | | | | |
|---|---|---|---|---|---|
| 275 | 81-85 | M | 17 | Acute | Fig. 3 |
| 276 | 41-45 | M | 44 | Convalescent | Fig. 3 |
| 277 | 81-85 | M | 13 | Acute | Fig. 3 |
| 278 | 36-40 | F | 51 | Convalescent | Fig. 3 |
| 279 | 61-70 | F | 53 | Convalescent | Fig. 3 |
| 280 | 36-40 | M | 45 | Convalescent | Both |
| 281 | 51-55 | M | 52 | Convalescent | Both |
| 282 | 46-50 | M | 9 | Acute | Both |
| 283 | 61-65 | M | 10 | Acute | Both |
| 284 | 56-60 | M | 69 | Convalescent | Both |
| 285 | 41-45 | M | 24 | Convalescent | Both |
| 286 | 81-85 | F | 11 | Acute | Both |
| 287 | 81-85 | F | 14 | Acute | Both |
| 288 | 81-85 | M | 5 | Acute | Both |
| 289 | 86-49 | M | 12 | Acute | Fig. 3 |
| 290 | 91-95 | F | 10 | Acute | Fig. 3 |
| 291 | 76-80 | M | 7 | Acute | Fig. 3 |
| 292 | 76-80 | M | 10 | Acute | Fig. 3 |
| 293 | 46-50 | M | 13 | Acute | Fig. 3 |
| 294 | 56-60 | M | 8 | Acute | Fig. 3 |
| 295 | 56-60 | M | 11 | Acute | Fig. 3 |
| 296 | 56-60 | F | 11 | Acute | Fig. 3 |
| 297 | 96-100 | F | 3 | Acute | Fig. 3 |
| 298 | 96-100 | F | 6 | Acute | Fig. 3 |
| 299 | 61-65 | M | 15 | Acute | Fig. 3 |
| 300 | 36-40 | M | 19 | Acute | Fig. 3 |
| 301 | 61-65 | F | 3 | Acute | Fig. 3 |
| 302 | 81-85 | F | 12 | Acute | Fig. 3 |
| 303 | 61-65 | F | 9 | Acute | Fig. 3 |
| 304 | 66-70 | M | 36 | Convalescent | Fig. 3 |
| 305 | 31-35 | F | 40 | Convalescent | Fig. 3 |
| 306 | 31-25 | F | 75 | Convalescent | Both |
| 307 | 41-35 | M | 86 | Convalescent | Both |
| 308 | 71-75 | F | 96 | Convalescent | Both |
| 309 | | | | Acute | Fig 1 |
| 400 | | | | Acute | Fig 1 |
| 401 | | | | Acute | Fig 1 |
| 402 | | | | Acute | Fig 1 |
| **Summary** | **Median =59** | **%M=61 %F=39** | **Mean. = 42d** | **%Acute=32 %Convalescent=68** | |

**Supplementary Table 3a.**

Bivariate and multivariable analysis examining potential independent associations between log-transformed saliva anti-Spike IgA.

| | **Bivariate analysis** | **Multivariable analysis** |
|---|---|---|
| **Predictor** | Beta coefficient (95% CI) | Beta coefficient (95% CI) |
| Age in years | 0.02 (0.01 to 0.04) | 0.02 (0.002 to 0.04) |
| Male sex | -0.4 (-1.1 to 0.3) | -0.4 (-1.1 to 0.3) |
| Time from vaccination to sample collection in days | 0.01 (-0.03 to 0.05) | 0.01 (-0.03 to 0.05) |
| Prior SARS-CoV-2 infection | 1.0 (0.4 to 1.5) | 0.9 (0.3 to 1.4) |

Abbreviations: CI = confidence interval

**Supplementary Table 3b.**

Bivariate and multivariable analysis examining potential independent associations between saliva anti-Spike IgG.

| | **Bivariate analysis** | **Multivariable analysis** |
|---|---|---|
| **Predictor** | Beta coefficient (95% CI) | Beta coefficient (95% CI) |
| Age in years | 0.6 (-0.1 to 1.2) | 0.4 (-0.2 to 1.1) |
| Male sex | -38.7 (-65.2 to -12.2) | -39.5 (-65.7 to -13.3) |
| Time from vaccination to sample collection | -0.9 (-4.7 to 2.8) | -0.4 (-1.9 to 1.1) |
| Prior SARS-CoV-2 infection | 21.3 (-0.3 to 42) | 19.7 (-1.5 to 41) |

Abbreviations: CI = confidence interval

**Supplementary Table 3c.**

Bivariate and multivariable analysis examining potential independent associations between log-transformed serum anti-RBD IgA.

| | **Bivariate analysis** | **Multivariable analysis** |
|---|---|---|
| **Predictor** | Beta coefficient (95% CI) | Beta coefficient (95% CI) |
| Age in years | 0.005 (-0.006 to 0.02) | 0.0008 (-0.01 to 0.01) |
| Male sex | 0.2 (-0.2 to 0.7) | 0.2 (-0.3 to 0.6) |
| Time from vaccination to sample collection | -0.04 (-0.06 to -0.01) | -0.04 (-0.06 to -0.01) |
| Prior SARS-CoV-2 infection | 0.5 (0.1 to 0.9) | 0.5 (0.2 to 0.9) |

Abbreviations: CI = confidence interval

**Supplementary Table 4.**
MSB-1 (longitudinal mRNA 2 doses cohort)

| Participant ID | Age | Sex | Vaccination Type |
|---|---|---|---|
| 403 | 20-25 | F | BNT162b2 |
| 404 | 26-30 | F | BNT162b2 |
| 405 | 26-30 | M | BNT162b2 |
| 406 | 20-25 | F | BNT162b2 |
| 407 | 20-25 | M | BNT162b2 |
| 408 | 20-25 | M | BNT162b2 |
| 409 | 20-25 | M | BNT162b2 |
| 410 | 41-45 | F | BNT162b2 |
| 411 | 20-25 | M | BNT162b2 |
| 412 | 20-25 | F | BNT162b2 |
| 413 | 31-35 | F | BNT162b2 |
| 414 | 36-40 | F | BNT162b2 |
| 415 | 56-50 | M | BNT162b2 |
| 416 | 56-60 | M | BNT162b2 |
| 417 | 20-25 | F | BNT162b2 |
| 418 | 36-40 | M | BNT162b2 |
| 419 | 20-25 | F | BNT162b2 |
| 420 | 20-25 | F | BNT162b2 |
| 421 | 31-35 | M | BNT162b2 |
| 422 | 26-30 | M | BNT162b2 |
| 423 | 26-30 | F | BNT162b2 |
| 424 | 26-30 | F | BNT162b2 |
| 425 | 26-30 | F | BNT162b2 |
| 426 | 20-25 | F | BNT162b2 |
| 427 | 26-30 | M | BNT162b2 |
| 428 | 46-50 | M | BNT162b2 |
| 429 | 41-45 | F | BNT162b2 |
| 430 | 41-45 | F | BNT162b2 |
| 431 | 26-30 | F | BNT162b2 |
| 432 | 26-30 | M | BNT162b2 |
| **Summary** | **Median age=27** | **%M=42 %F=56** | |

**Supplementary Table 5.**
Long Term Care (LTC) residents outbreak cohort

| Participant ID | Age | Sex | Vaccination type | PCR-confirmed status at time of outbreak |
|---|---|---|---|---|
| 433 | 91-95 | Male | mRNA-1273 | Infected |
| 434 | 91-95 | Female | mRNA-1273 | Not infected |
| 435 | 91-95 | Male | mRNA-1273 | Not infected |
| 436 | 96-100 | Female | mRNA-1273 | Infected |
| 437 | 91-95 | Female | mRNA-1273 | Not infected |
| 438 | 96-100 | Female | mRNA-1273 | Not infected |
| 439 | 81-85 | Female | mRNA-1273 | Not infected |
| 440 | 86-90 | Female | mRNA-1273 | Not infected |
| 441 | 91-95 | Female | mRNA-1273 | Not infected |
| 442 | 91-95 | Female | mRNA-1273 | Not infected |
| 443 | 91-95 | Male | mRNA-1273 | Not infected |
| 444 | 76-80 | Male | mRNA-1273 | Not infected |
| 445 | 91-95 | Male | mRNA-1273 | Infected |
| 446 | 91-95 | Female | mRNA-1273 | Infected |
| 447 | 86-90 | Male | mRNA-1273 | Infected |
| 448 | 96-100 | Female | mRNA-1273 | Not infected |

**Supplementary Table 6.**
Sheba Medical Centre Cohort (breakthrough infection post-vaccination)

| Participant ID | Participant Age | Participant Sex | PCR-confirmed status |
|---|---|---|---|
| 449 | 46-50 | F | uninfected |
| 450 | 46-50 | F | uninfected |
| 451 | 36-40 | M | infected |
| 452 | 46-50 | F | uninfected |
| 453 | 36-40 | F | uninfected |
| 454 | 41-45 | M | uninfected |
| 455 | 51-55 | F | uninfected |
| 456 | 46-50 | F | uninfected |
| 457 | 46-50 | M | uninfected |
| 458 | 41-45 | M | uninfected |
| 459 | 51-55 | M | uninfected |
| 460 | 46-50 | F | uninfected |
| 461 | 51-55 | M | infected |
| 462 | 51-55 | M | uninfected |
| 463 | 46-50 | F | uninfected |
| 464 | 46-50 | F | uninfected |
| 465 | 41-45 | M | uninfected |
| 466 | 41-45 | F | uninfected |
| 467 | 46-50 | M | uninfected |
| 468 | 56-60 | F | uninfected |
| 469 | 51-55 | F | uninfected |
| 470 | 46-50 | F | uninfected |
| 471 | 41-45 | F | uninfected |
| 472 | 46-50 | M | uninfected |
| 473 | 46-50 | F | uninfected |
| 474 | 41-45 | M | uninfected |
| 475 | 61-65 | F | uninfected |
| 476 | 61-65 | M | uninfected |
| 477 | 41-45 | F | infected |
| 478 | 31-35 | M | infected |
| 479 | 61-65 | F | uninfected |
| 480 | 51-55 | F | uninfected |
| 481 | 36-40 | M | uninfected |
| 482 | 41-45 | M | infected |
| 483 | 26-30 | M | uninfected |
| 484 | 46-50 | M | uninfected |

| | | | |
|---|---|---|---|
| 485 | 41-45 | M | uninfected |
| 486 | 41-45 | M | uninfected |
| 487 | 46-50 | F | uninfected |
| 488 | 31-35 | M | uninfected |
| 489 | 51-55 | F | uninfected |
| 490 | 46-50 | F | uninfected |
| 491 | 51-55 | M | uninfected |
| 492 | 36-40 | M | uninfected |
| 493 | 46-50 | M | uninfected |
| 494 | 36-40 | M | uninfected |
| 495 | 46-50 | M | uninfected |
| 496 | 46-50 | M | uninfected |
| 497 | 51-55 | F | infected |
| 498 | 46-50 | M | infected |
| 499 | 41-45 | F | uninfected |
| 500 | 46-50 | F | infected |
| 501 | 46-50 | F | uninfected |
| 502 | 46-50 | F | uninfected |
| 503 | 51-55 | F | uninfected |
| 504 | 46-50 | F | infected |
| 505 | 41-45 | M | uninfected |
| 506 | 41-45 | F | uninfected |
| 507 | 46-50 | F | uninfected |
| 508 | 46-50 | F | uninfected |
| 509 | 46-50 | F | uninfected |
| 510 | 61-65 | F | uninfected |
| 511 | 36-40 | M | infected |
| 512 | 41-45 | M | uninfected |
| 513 | 46-50 | F | uninfected |
| 514 | 46-50 | F | infected |
| 515 | 36-40 | M | uninfected |
| **Summary** | **Median Age= 47** | **%F=54%**<br>**%M=46%** | |
